# Polygenic risk scores enhance the identification of carriers of monogenic forms of idiopathic pulmonary fibrosis

**DOI:** 10.64898/2026.04.16.26350967

**Authors:** Aitana Alonso-González, David Jáspez, José M. Lorenzo-Salazar, Andrea Delgado, Ariadna Quintero-Bacallado, Shwu-Fan Ma, Emma Strickland, Josyf Mychaleckyj, John S. Kim, Yong Huang, Ayodeji Adegunsoye, Justin M. Oldham, Toby M. Maher, Beatriz Guillén-Guio, Louise V. Wain, Richard J. Allen, Gauri Saini, R. Gisli Jenkins, Maria Molina-Molina, David Zhang, Christine Kim Garcia, Fernando J. Martinez, Imre Noth, Carlos Flores

**Affiliations:** Research Unit, Hospital Universitario Nuestra Señora de Candelaria, Instituto de Investigación Sanitaria de Canarias (IISC), Santa Cruz de Tenerife, Spain; Genomics Division, Instituto Tecnológico y de Energías Renovables, Santa Cruz de Tenerife, Spain; Division of Pulmonary and Critical Care Medicine, University of Virginia, Charlottesville, VA USA; Center for Public Health Genomics; University of Virginia, Charlottesville, VA, USA; Section of Pulmonary and Critical Care Medicine, University of Chicago, Chicago, IL USA; Division of Pulmonary and Critical Care Medicine, University of Michigan, Ann Arbor, MI USA; National Heart and Lung Institute, Imperial National Institute of Health and Care Research, Biomedical Research Centre, Imperial College London, London, UK; Division of Pulmonary and Critical Care Medicine, University of Southern California, Los Angeles, CA USA; Division of Public Health and Epidemiology, University of Leicester, Leicester, UK; University Hospitals of Leicester NHS Trust, Leicester, UK; CIBER de Enfermedades Respiratorias (CIBERES), Instituto de Salud Carlos III, Madrid, Spain; Royal Papworth University Hospital NHS Foundation Trust and University of Nottingham, UK; Servei de Pneumologia, Laboratori de Pneumologia Experimental, IDIBELL, Barcelona, Spain; Campus de Bellvitge, Universitat de Barcelona, Barcelona, Spain; Department of Medicine, Columbia University Irving Medical Center, New York, NY, USA; Columbia Precision Medicine Initiative, Columbia University Irving Medical Center, New York, NY, USA; Weill Cornell Medical Center, New York, NY USA; Facultad de Ciencias de la Salud, Universidad Fernando Pessoa Canarias, Las Palmas de Gran Canaria, Spain

**Keywords:** Idiopathic pulmonary fibrosis, polygenic risk score, monogenic variants

## Abstract

**Background:** Idiopathic pulmonary fibrosis (IPF) is a rare disease with a poor prognosis. Disease risk involves rare and common genetic variants. However, an inverse association have been described between them. Accordingly, IPF patients with a higher polygenic risk score (PRS) for IPF are less likely to carry rare deleterious variants and vice versa. Here, we evaluate weather PRS of IPF could serve as an additional criterion to patient prioritisation for rare variant discovery.

**Methods:** We identified carriers based on the presence of rare qualifying variants (QVs) in genes linked to monogenic forms of pulmonary fibrosis in 888 IPF patients from the Pulmonary Fibrosis Foundation Patient Registry (PFF-PR). Genome-wide association study (GWAS) summary statistics from independent cohorts were used to construct a whole-genome PRS (WG-PRS) using a clumping and thresholding method (C+T) and a Bayesian method (SBayesRC). PRS were also derived from 19 known common sentinel IPF variants (Sentinel-PRS). Logistic regression models were used to evaluate associations between PRS and carrier status. Discriminatory performance was evaluated using area under the curve (AUC) analysis, and comparisons were made with DeLong’s test. Validation was performed in 472 IPF individuals from the UK PROFILE cohort.

**Results:** IPF-PRS were strongly associated with the QVs carrier status: Odds Ratio [OR] 0.65 (95% Confidence Interval [CI] 0.53-0.79) for WG-PRS_C+T_, OR 0.71 (95% CI 0.59-0.86) for WG-PRS_SBayesRC_, and OR 0.77 (95% CI 0.63-0.94) for Sentinel-PRS. Adding WG-PRS to the patient’s personal clinical history improved the prediction of QVs carriers: AUC=0.62 for the clinical model, AUC=0.68 for WG-PRS_C+T_ (DeLong’s test, p=9.54×10^-4^) and AUC=0.66 for WG-PRS_SBayesRC_ (DeLong’s test, p=0.02). Adding of IPF-PRS to clinical variables correctly reclassified 22.8% of carriers when using WG-PRS_C+T_, 20.8% when using Sentinel-PRS, and 16.7% for WG-PRS_SBayesRC_. WG-PRS_SBayesRC_ and the Sentinel-PRS also demonstrated improved prediction of QVs carriers in telomere-related genes in PROFILE.

**Conclusions:** Incorporating IPF-PRS into a model based on the patient’s clinical history improves the identification of QVs carriers. Although the overall discriminatory power was moderate, these findings raise de the possibility of using WG-PRS as useful criterion for rare variant discovery in patients with IPF and enhance decision-making.

## BACKGROUND

Idiopathic pulmonary fibrosis (IPF) is a rare complex late-onset disease characterized by progressive and irreversible lung scarring of the lung parenchyma (1). Genetic factors play a significant role in the disease aetiology, with common and rare variants increasing disease risk. Genome-wide association studies (GWAS) of sporadic IPF have identified several genetic loci, most notably a variant (rs35705950-T) in the promoter region of the *MUC5B* gene with an odds ratios (OR) greater than 5 (2–5). Next-generation sequencing (NGS) studies including familial forms of pulmonary fibrosis (PF) have led to the identification of rare deleterious variants (RDVs) associated with the disease (6–8). These variants cluster principally in genes involved in telomere biology, surfactant metabolism, and mitotic spindle assembly.

While the median survival of IPF patients is 3-5 years after diagnosis (9), the clinical course varies significantly among patients. The reason for this is not fully understood, although genetics may partially account for such heterogeneity. Recent studies suggest that RDVs, particularly those in telomere-related genes, may define a distinct subtype associated with poorer survival and extrapulmonary complications (10,11). Therefore, identification of these variants in clinical practice is critical, given their implications for risk stratification, treatment decision-making, and early identification of high-risk relatives.

In an effort to improve the identification of patients that could benefit from genetic testing, the European Respiratory Society recently published a statement emphasizing the need for genetic evaluation in individuals with a family history of PF, clinical suspicion of a telomere syndrome, or early-onset disease (12). However, some IPFpatients carrying RDVs do not meet any of these criteria and therefore remain uncharacterized genetically. For instance, at least 10% of patients with clinically sporadic IPF carry RDVs and may therefore represent unrecognized familial cases. In addition, because early symptoms are often nonspecific, diagnosis frequently occurs after the age of 68, even among individuals with a genetic predisposition (11).

Recent studies have proposed the use of polygenic risk scores (PRS) as a cost-effective tool for prioritizing patients for rare variant screening (13). This approach is supported by the observation that individuals with a disease but low polygenic burden are more likely to harbour rare large effect variants (14). This inverse relationship between polygenic risk and rare variant carrier status has been described across a range of complex disorders such as prostate cancer or hypertrophic cardiomyopathy (15,16). However, the magnitude of this relationship, and therefore, the utility of this approach, is expected to vary depending on the disease genetic architecture.

We recently described that, among IPF patients, PRS of IPF (IPF-PRS) constructed from 19 sentinel GWAS variants (Sentinel-PRS) were inversely associated with the prevalence of rare variants affecting monogenic adult-onset PF genes (11), suggesting an independent contribution to disease. Accordingly, patients with higher PRS may be less likely to harbour a RDV, while those with lower PRS may be more likely to carry one. This finding supports the idea that the polygenic component of the disease could be leveraged to stratify PF patients and improve genetic testing results. Specially, in the absence of an extensive family history information or a diagnosis delay. However, that study did not explore the predictive performance of the PRS for distinguishing rare variant carriers from non-carriers, nor did it evaluate a whole-genome model for PRS estimates (WG-PRS).

Here, we first examined the association between WG-PRS and Sentinel-PRS with rare variants with predicted deleterious potential. Then, we assessed whether incorporating PRS into models based on the patient’s clinical history currently used in practice may improve the identification of those that are carriers of rare variants affecting monogenic adult-onset PF genes.

## MATERIAL AND METHODS

### Study design

The main focus of the study was to assess whether different models of IPF-PRS estimation could optimize the study of potential carriers of rare variants in PF genes that are expected to be causal of disease. To this end, we first derived IPF-PRS using both WG-PRS and Sentinel-PRS based on published IPF GWAS results (4) to evaluate their association with the rare variants previously prioritized in genetic data from two cohorts of IPF patients (11). We then evaluated the performance of the different IPF-PRS estimates combined with patient’s clinical history in predicting carriers in comparison with the performance of the standard model based only on the patient’s clinical history and whether the models including the IPF-PRS estimates could identify additional carriers. In the first stage, we used whole-genome sequencing (WGS) data from the Pulmonary Foundation Patients Registry (PFF-PR). WGS data from the Prospective Observation of Fibrosis in the Lung Clinical Endpoints (PROFILE) cohort was used for validation of findings.

### Study cohorts

#### The PFF-PR cohort

We included 888 unrelated patients diagnosed with IPF from the PFF-PR which have been previously described (11). Data from all of them were retained after individual-level quality control (QC) assessments were conducted. Briefly, The PFF-PR is a multicentre, observational study in the USA which collects baseline and longitudinal demographic and clinical information about patients with interstitial lung disease (ILD). In addition, blood samples are obtained to study molecular markers of the onset or progression of disease (17). WGS of the entire cohort was performed at Psomagen (Rockville, MD, USA) and Illumina DRAGEN Bio-IT Platform Germline Pipeline v3.10.4 (Illumina Inc.) was used for obtaining variant calling by aligning to the GRCh38 reference genome. TelSeq 0.0.2 was used to estimate age-unadjusted telomere length (TL) measures from WGS alignment files.

#### The PROFILE cohort

For validation, we used data from 472 patients diagnosed with IPF from PROFILE passing the individual-level QC assessments (11). The PROFILE cohort is a UK, prospective, multicentre, longitudinal study which includes in total 541 patients with fibrotic ILD. Patients were followed for disease progression for three years and blood samples were also collected (18). WGS was obtained at Human Longevity (San Diego, CA, USA). Variant calling was performed with DRAGEN Bio-IT Platform Germline Pipeline v3.0.7, using the GRCh38 as the reference genome. TelSeq 0.0.2 was then applied to estimate age-unadjusted TL measures from WGS alignment files. Both studies were conducted according to The Declaration of Helsinki and written informed consent was obtained from all participants. Ethical approval for the PFF-PR was granted by the Institutional Review Board from the University of Michigan (MI, USA; ethics reference number HUM00111461). Ethical approval for the PROFILE study was granted by the Royal Free Hospital Research Ethics Committee (London, UK; ethics reference number 10/H0720/12) and University of Northampton Research Ethics Committee (Northampton, UK; ethics reference number 10/H0402/2).

### Qualifying variants and carrier definition

We defined qualifying variants (QVs) as a proxy for RDVs based on predefined filtering criteria. In the absence of functional validation or co-segregation data, we classified these variants using *in silico* predictions and established pathogenicity frameworks, representing them as putative deleterious variants potentially affecting protein function. There were identified from WGS data from patients of the two cohorts. Briefly, the QVs were identified in 13 PF genes (*TERC, TERT, TINF2, DKC1, RTEL1, PARN, NAF1, ZCCHC8, SFTPC, SFTPA1, SFTPA2, SPDL1*, and *KIF15*). As described elsewhere (11), they will be referred for simplicity as monogenic adult-onset PF genes. QVs were obtained after filtering out variants based on read depth <10, mapping quality <50, fraction of missing genotypes >0.05, and global allele frequency (AF)>0.0005 in gnomAD v2.1. Then, protein-truncating variants and missense variants with a CADD>15 were retained. For the non-coding RNA gene *TERC*, variants with AF<0.0005 and annotated by ClinVar as pathogenic (P), likely pathogenic (LP), or of uncertain significance (VUS) were retained. Further details regarding the selection of genes and the bioinformatic filters applied are described elsewhere (11). For the purpose of this study, we defined the carrier status by the presence of at least one of these QVs.

### Estimation of polygenic risk scores of idiopathic pulmonary fibrosis

#### Specific quality control of WGS data

Single nucleotide polymorphisms (SNPs) with a minor AF <0.01 were excluded from WGS data using BCFtools v.1.12 (https://samtools.github.io/bcftools/bcftools.html). Genotyping QC was then performed on the remaining SNPs using PLINK v2.00a3.6. SNPs were excluded if deviating largely from Hardy-Weinberg equilibrium expectations (p<1×10^-6^), call rate <95%, or if they were in sexual chromosomes or the mitogenome.

#### Principal components of genetic heterogeneity

After QC of WGS data, principal components (PCs) were calculated using PLINK v2.00a3.6. To this aim, regions with known high linkage disequilibrium (LD) were first removed. Subsequently, LD pruning (indep-pairwise 100 5 0.012) was performed in order to eliminate SNPs in high LD in a given window. Finally, PCs were estimated on the remaining 101,588 independent SNPs in PFF-PR, and 143,214 independent SNPs in PROFILE. As the first two PCs explain most of the genetic variation, they were used as covariates in the subsequent analyses to account for population heterogeneity.

#### PRS modelling

GWAS summary statistics from a large IPF meta-GWAS study (4) were used as the base dataset to estimate IPF-PRS in patients from the PFF-PR and PROFILE (target datasets). Sentinel-PRS were derived based on the 19 genome-wide significant IPF risk variants. WG-PRS were constructed using two broadly used methods which differ in variant selection: i) clumping and thresholding (WG-PRS_C+T_), and ii) a Bayesian method (WG-PRS_SBayesRC_).

WG-PRS_C+T_ were estimated using PRSice-2 (19). Briefly, clumping is performed to remove SNPs in high LD and then patient’s PRS are derived using only those SNPs with a GWAS association p-value below a certain threshold. The following thresholds were used: 1×10^-8^, 5×10^-8^, 1×10^-7^, 5×10^-7^, 1×10^-6^, 5×10^-6^, 1×10^-5^, 5×10^-5^, 1×10^-4^, 5×10^-4^, 0.05, 0.5, and 1. The significance threshold for the WG-PRS_C+T_ was set at 1×10^-3^ to correct for the multiple comparisons.

WG-PRS_SBayesRC_ were estimated using SBayesRC (20). Briefly, SBayesRC adjusts effect size estimates of all SNPs (approximately 7 million common variants) using a Bayesian approach which incorporates functional annotation and LD information. Individuals IPF-PRS were then derived as the sum of risk alleles from the IPF GWAS variants weighted by their effect sizes:

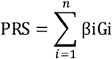

where βi is the OR from variant i as identified in the IPF meta-GWAS study (4), Gi represents the number of risk alleles carried at the variant i, and n represents the conditionally independent signals identified elsewhere. Results were then standardized as z-scores using the following formulae:

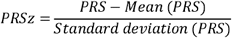

### Clinical variables from patient’s clinical history

To construct a clinical model aimed at prioritizing carriers of RDVs, we selected three variables commonly used in standard clinical practice: age at disease onset, TL, and family history. Early-onset disease was defined as a diagnosis at or before 50 years of age. Severe telomere shortening was defined as TL values within the lowest 10th percentile of each cohort. A positive family history was defined as the known presence of at least one first-degree relative affected by ILD; all other cases were considered sporadic. Information on family history was not available for the PROFILE cohort.

### Statistical analysis

Logistic regression models were used to assess the association of IPF-PRS (independently for all three models) and clinical variables (age at disease onset, TL, and family history) with the carrier status. PRS models were adjusted for the two first PCs and clinical variables.

To evaluate predictive performance of each model, we estimated the area under the curve (AUC) and the receiver operating characteristic (ROC) curve. The AUC of models including IPF-PRS as well as the patient’s personal clinical history was compared to the AUC of a model with the patient’s personal clinical history alone using the DeLong’s test. Specificity, sensitivity, positive predictive value, negative predictive value, true positives, true negatives, false positives, and false negatives were estimated for each model considering a risk threshold of 5%, 10%, 15%, and 20%. Calibration of each model was evaluated comparing predicted and observed values using the Hosmer-Lemeshow test.

To assess the improvement of prediction performance gained by adding IPF-PRS estimates to the patient’s personal clinical history alone model we used reclassification tables and the net reclassification index (NRI) for each risk threshold (5%, 10%, 15%, and 20%). NRI was calculated separately for cases (QVs carriers), NRI_C_, and controls (non-carriers), NRI_nc_. NRI_c_ was estimated as the proportion of cases correctly reclassified as high risk by the IPF-PRS with the patient’s personal clinical history model minus the proportion of cases incorrectly reclassified as low risk. Similarly, NRI_nc_ was estimated as the proportion of non-cases correctly reclassified as low risk by the IPF-PRS with the patient’s personal clinical history model minus the proportion of non-cases incorrectly reclassified as high risk.

Decision curve analysis (DCA) was also used to evaluate whether adding IPF-PRS estimates to the patient’s personal clinical history could help prioritize patients for gene sequencing. We plotted the net benefit of each model (clinical history alone and IPF-PRS plus clinical history) across a range of risk thresholds and compared them with the “sequence all” and “sequence none” strategies.

Statistical analyses were performed with R v. 4.3.1 (21), with p-values<0.05 considered significant. The *pROC* package v.1.18.5 was used for visualizing ROC curves and for comparing AUCs. *PredictABEL* v.1.2-4 was used to estimate reclassification tables. The *dcurves* package v.0.5.0 was used for DCA analysis. The *probably* package v.1.2.0 and the *ResourceSelection* package v.0.3-6 were used to assess model calibration by plotting the relationship between predicted and observed values and by performing calibration tests.

### Sensitivity analysis

To ensure that the association of WG-PRS with the carrier status was not driven by the *MUC5B* risk variant, IPF-PRS were also estimated after excluding variants in the ±500 kb flanking regions of rs35705950. To assess potential bias associated with the definition of the QVs, alternative and stricter definitions of QVs were used in the analysis (**Additional file 1: Supplementary Table 1**). This also included a category of rare synonymous variants in the 13 monogenic adult-onset PF genes, which were expected to represent neutral variation and were therefore used as a null model in the association analyses.

## RESULTS

### Characteristics of patients with IPF from the PFF-PR

The discovery cohort included 888 PFF-PR IPF patients as described elsewhere (11), where 144 carriers of QVs were identified (16.2%). The PROFILE cohort included 472 IPF individuals, of whom 52 were QVs carriers (11.0%) (11). In the PFF-PR we found a robust association between the carrier status and increased risk of severely reduced of TL (OR=3.16; 95% Confidence Interval [CI]=1.91-5.15; p=4.79×10^-6^), family history of ILD (OR=2.27; 95% CI=1.50-3.39; p=7.83×10^-5^), and risk of age of onset before 50 years old (OR=3.33; 95% CI=1.29-8.20; p=0.01). In contrast, in PROFILE neither age at diagnosis before 50 years nor severe telomere shortening was associated with overall QV carrier status; family history data were not available. However, when restricting the analysis to carriers of variants in telomere-related genes, severe telomere shortening was associated with carrier status (OR=2.28; 95% CI=1.00–4.81; p=0.04).

The number of QVs carriers stratified by these three variables is detailed in Table 1.

**Table 1.**
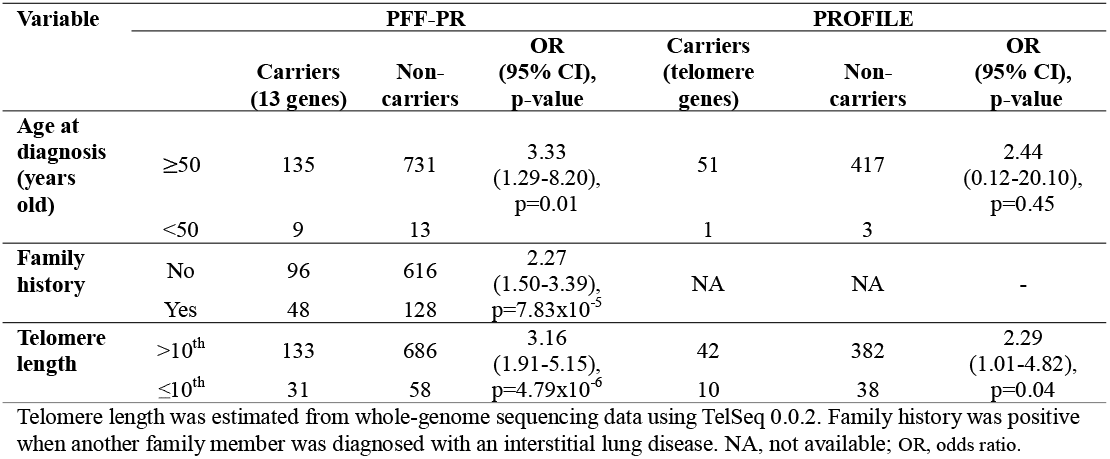
Carriers and non-carriers of qualifying variants stratified by clinical variables in the PFF-PR and PROFILE cohorts.

### Associations of IPF-PRS with the QVs carrier status

The best performing WG-PRS_C+T_ model was set at p-value threshold of 5×10^-4^ and included 1,097 genetic variants in PFF-PR and 1,114 in PROFILE (**Additional file 1: Supplementary Figure 1**).

In the discovery cohort, WG-PRS_C+T_ were strongly associated with the carrier status, with an OR of 0.65 (95% CI=0.53-0.79; p=1.98×10^-5^) (**Table 2**). When WG-PRS_C+T_ were divided into quintiles, the highest quintile was associated with substantially lower odds of being a carrier compared to the lowest quintile (OR=0.16; 95% CI=0.08-0.35; p=2.87×10^-6^) (**Figure 1A**). WG-PRS_SBayesRC_ further supported the robustness of these associations (OR=0.71; 95% CI=0.59-0.86; p=4.47×10^-4^) (**Table 2**). In addition, their stratification in quintiles also revealed a lower proportion of carriers in the highest quintile in comparison to the lowest quintile (OR=0.46; 95% CI=0.26-0.82; p=8.86×10^-3^) (**Figure 1A**). Sentinel-PRS also showed a significant association with the carrier status, although the strength of the association was attenuated (OR=0.77; 95% CI=0.63-0.94, p=0.013) and the differences between the first and last quintiles were not significant (**Table 2, Figure 1A**). In the validation cohort (PROFILE), no association was observed between overall QV carrier status and any IPF-PRS model, consistent with the absence of association between clinical predictors and carrier status in this cohort. However, when restricting the analysis to carriers of telomere-related gene variants, the direction and magnitude of effect were comparable to those observed in PFF-PR. Both Sentinel-PRS (OR=0.68; 95% CI=0.50–0.92; p=0.01) and WG-PRS_SBayesRC_ (OR=0.68; 95% CI=0.50–0.92; p=0.01) were significantly associated with telomere-gene carrier status (**Table 2**), and quintile stratification similarly showed a lower odds of being a carrier in the highest PRS quintile compared to the lowest quintile (**Figure 1B**).

**Figure 1.**
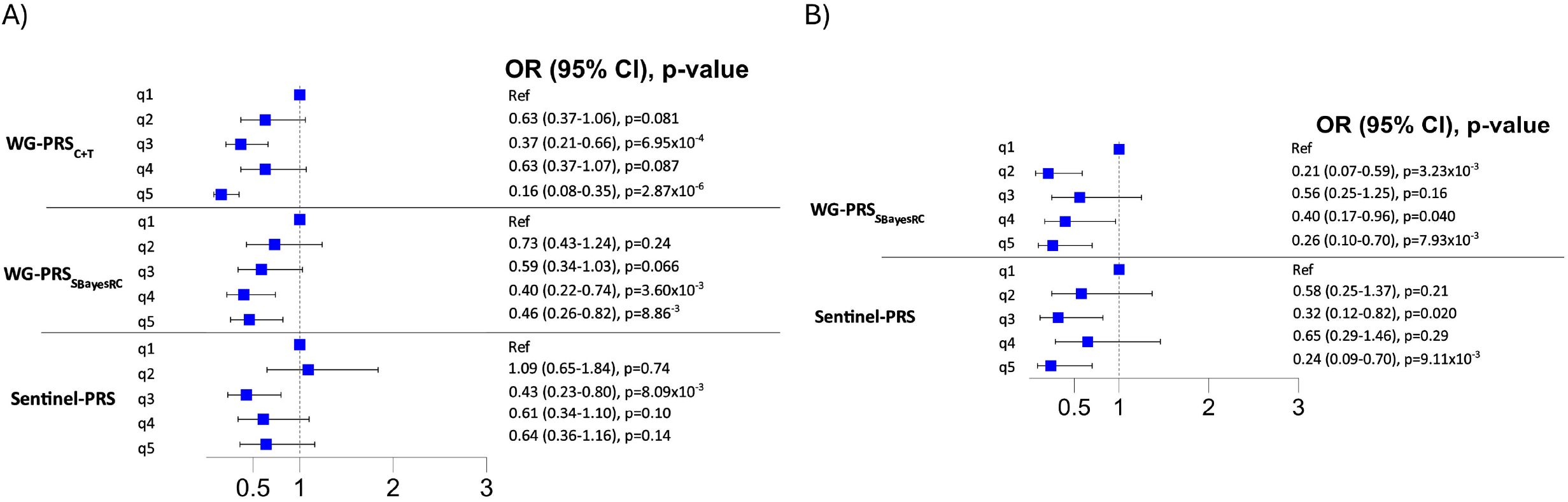
Forest plot by quintile showing odds ratios (OR) and the 95% confidence intervals (CI) of the IPF-PRS association with the qualifying variant carrier status for each quintile (fifth quintile used as the refence group). Models were adjusted for the patient’s personal clinical history data. A) PFF-PR. B) PROFILE.

**Table 2.** Association of IPF-PRS with the qualifying variant carrier status in the PFF-PR and PROFILE cohorts.

| IPF cohort | PRS model | SNPs included | Model threshold p-value | Covariates | OR (95% CI) | p-value |
| --- | --- | --- | --- | --- | --- | --- |
| PFF-PR* | WG-PRS<sub>C+T</sub> | 1097 | 5x10<sup>-4</sup> | Age<50, FPF, TL≤10<sup>th</sup>, PC1-2 | 0.65 (0.53-0.79) | 1.98x10<sup>-5</sup> |
|  | WG-PRS<sub>SBayesRC</sub> | ~7 million | - |  | 0.71 (0.59-0.86) | 4.47x10<sup>-4</sup> |
|  | Sentinel-PRS | 19 | 1x10<sup>-8</sup> |  | 0.77 (0.63-0.94) | 0.01 |
|  | WG-PRS<sub>C+T</sub> without *MUC5B* | 1055 | 5x10<sup>-4</sup> |  | 0.72 (0.59-0.88) | 1x10<sup>-3</sup> |
|  | WG-PRS<sub>SBayesRC</sub> without *MUC5B* | ~7 million | - |  | 0.83 (0.69-1.00) | 0.05 |
| PROFILE# | WG-PRS<sub>C+T</sub> | 1114 | 5x10<sup>-4</sup> | Age<50, TL≤10<sup>th</sup>, PC1-2 | 0.87 (0.64-1.18) | 0.40 |
|  | WG-PRS<sub>SBayesRC</sub> | ~7 million | - |  | 0.68 (0.50-0.92) | 0.01 |
|  | Sentinel-PRS | 19 | 1x10<sup>-8</sup> |  | 0.68 (0.50-0.92) | 0.01 |

\*Carriers were defined as patients with at least one QVs in telomere or not telomere genes; #Carriers were defined as patients with at least one QVs in telomere genes. OR, odds ratio; CI, confidence interval; IPF-PRS, polygenic risk score of idiopathic pulmonary fibrosis; TL, telomere length; FPF, familial history of interstitial lung disease; PC1-2, principal components 1 and 2.

Sensitivity analyses were performed in the PFF-PR. First, the exclusion of the MUC5B locus from the WG-PRS_C+T_ modelling attenuated the association between IPF-PRS and the QVs carrier status, although it remained statistically significant (OR=0.72; 95% CI=0.59-0.88; p=1×10^-3^) (**Table 2**). Second, we tested the association of IPF-PRS with alternative definitions of QVs to increase the likelihood of them being potentially RDVs (**Additional file 1: Supplementary Table 1**). Interestingly, the carrier status remained consistently associated with IPF-PRS across the three models. Reassuringly, the null model testing the presence of rare synonymous variants showed no association with any of the IPF-PRS models (**Additional file 1: Supplementary Table 2**).

### Performance of IPF-PRS models and patient’s clinical history

We next explored whether IPF-PRS may improve prediction models for QVs carrier status currently recommended based on the age at diagnosis before 50 years old, family history, and TL.

Our results in the PFF-PR cohort showed that integrating WG-PRS or Sentinel-PRS to these clinical findings performed better than a model including only the patient’s clinical history data or a model including only the IPF-PRS (**Table 3, Figure 2A, Additional file 1: Supplementary Figure 2, Supplementary Figure 3, Supplementary Table 3**). However, only WG-PRS yielded a statistically significant improvement. The model combining WG-PRS_C+T_ with the patient’s clinical history achieved an AUC of 0.68 compared with 0.62 for the patient’s clinical data-only model (p=9.54×10^-4^). The predictive ability of a model incorporating WG-PRS_SBayesRC_ showed a similar trend (AUC=0.66 vs AUC=0.62; p-value=0.02) (**Table 3, Figure 2A**). In PROFILE, the predictive ability of models including IPF-PRS alone, and of those integrating IPF-PRS with the patient’s clinical history was higher than a model including only the patient’s clinical history (AUC=0.64 for Sentinel-PRS; AUC=0.63 for WG-PRS_SBayesRC_; vs. AUC=0.58 for model with clinical data alone) (**Table 3, Figure 2B, Additional file 1: Supplementary Figure 3, Supplementary Table 4**).

**Table 3.**
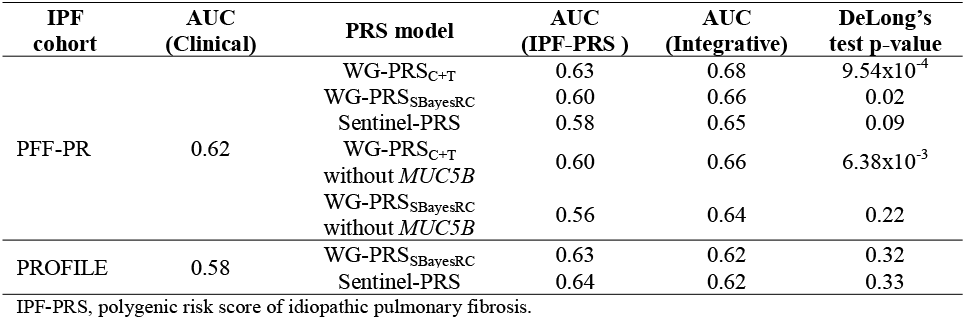
Area under the curve (AUC) for carrier prediction among patients with idiopathic pulmonary fibrosis. DeLong’s test p-values were used to compare models integrating the IPF-PRS with the patient’s clinical history (integrative) against the model with the patient’s clinical history only (clinical). Results for the model including only the IPF-PRS estimates are included for reference.

**Figure 2.**
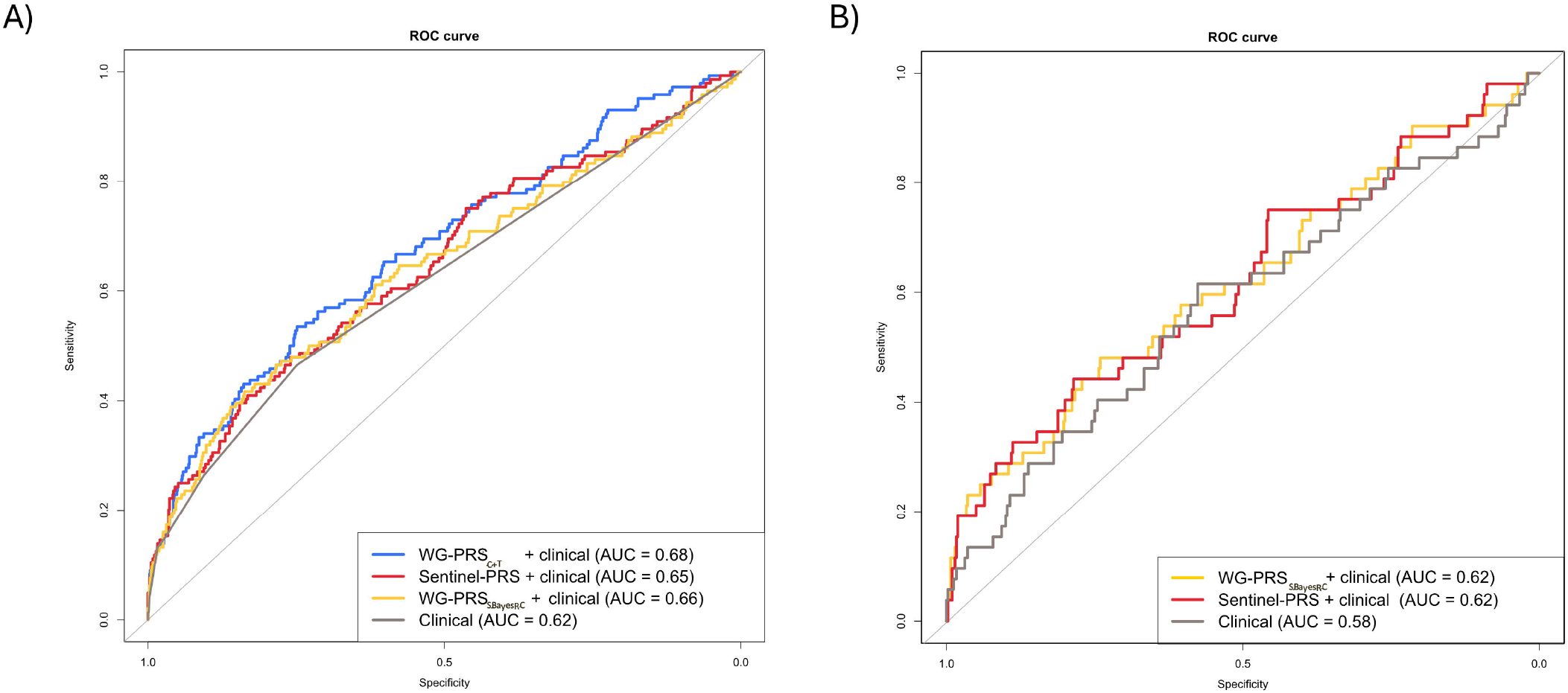
Receiver operating characteristic (ROC) curves for carrier prediction in IPF patients based on patient’s clinical history (age of diagnosis, family history, and telomere length) and models also integrating PRS-IPF. A) PFF-PR. B) PROFILE.

We also evaluated the performance of models after stratifying by clinical variables in both cohorts. In the PFF-PR Integrating the WG-PRS_C+T_ with the patient’s clinical history showed a better discriminatory power among patients with short TL (AUC=0.75; p value=0.04), and in patients with family history (AUC=0.75; p-value=0.02). This improvement was also observed for models alternatively integrating Sentinel-PRS and WG-PRS_SBayesRC_ albeit with non-significant differences between the categories (**Additional file 1: Supplementary Table 5**). In PROFILE we also observed a better discriminatory power of IPF-PRS among individuals with short TL for both IPF-PRS estimators (**Additional file 1: Supplementary Table 6**).

To assess robustness, sensitivity analyses were conducted in the discovery cohort. When alternative QV definitions were applied, models integrating the IPF-PRS along with the patient’s clinical history also performed better than models with the clinical history alone (**Additional file 1: Supplementary Figure 4, Supplementary Table 7**). The predictive ability further improved when analyses were restricted to variants with more deleterious potential (**Additional file 1: Supplementary Figure 4**). In this case, the differences with results of a model based on patient’s clinical history alone were statistically significant for all but the rare protein-truncating variant (PTV) category (**Additional file 1: Supplementary Table 7**). By contrast, for rare synonymous variants, which typically represent neutral variation, neither the clinical model nor the PRS model showed higher AUCs (**Additional file 1: Supplementary Table 7**). Finally, WG-PRS excluding the *MUC5B* locus also supported these results, showing that integrating the WG-PRS estimates with the clinical data in the prediction models improve the identification of QVs carriers and that such improvement did not depend solely on the *MUC5B* effect (**Table 3, Additional file 1: Supplementary Figure 2**).

### Improvement in carrier identification by adding PRS to clinical variables

In the PFF-PR, using reclassification tables upon a threshold of 15%, the models integrating IPF-PRS with the patient’s clinical history correctly reclassified 22.2% of QVs carriers when using WG-PRS_C+T_, 20.8% when using Sentinel-PRS, and 16.7% for WG-PRS_SBayesRC_ (**Figure 3A, Additional file 1: Supplementary Figure 5, Supplementary Figure 6, Supplementary Figure 7**). In PROFILE, at the same 15% threshold, integrating IPF-PRS with clinical variables resulted in reclassification of 9.61% of carriers using WG-PRS_SBayesRC_ and 19.23% using Sentinel-PRS (**Figure 3B, Additional file 1: Supplementary Figure 8**).

**Figure 3.**
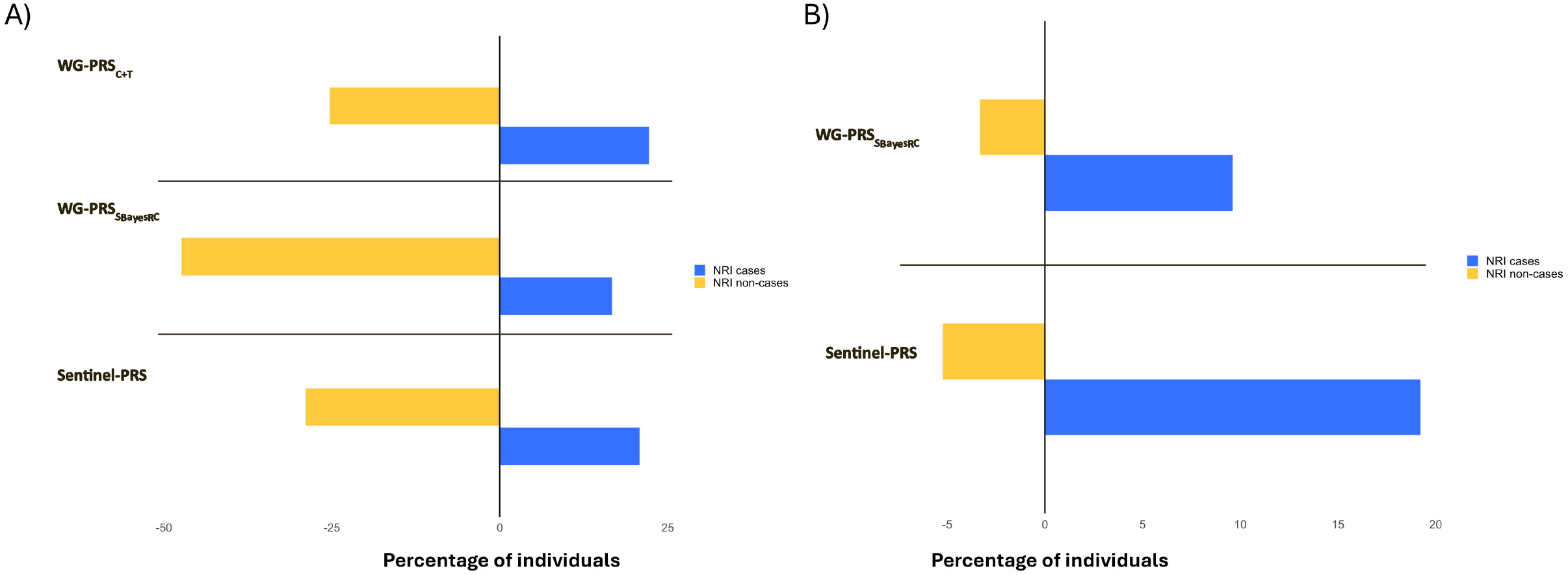
Visual representation of the net reclassification of patients comparing the models integrating the IPF-PRS with the patient’s clinical history vs. the model only including the patient’s clinical history using a risk threshold of 15%. A) PFF-PR. B) PROFILE.

In the PPFPR the use of alternative predefined risk at 5% did not show any improvement in the reclassification of cases nor of controls. By contrast, integrating the IPF-PRS with the patient’s clinical history at thresholds of 10% and 20% showed an improvement in the reclassification of controls for WG-PRS and Sentinel-PRS, although not for cases (**Additional file 1: Supplementary Figure 5, Supplementary Figure 6, Supplementary Figure 7**). DCA assessments in the discovery cohort, showed that integrating the IPF-PRS with patient’s clinical data did not result in higher net benefits compared with strategies based solely on patient’s clinical data alone or alternatively to perform gene sequencing in all IPF patients (**Additional file 1: Supplementary Figure 9, Supplementary Table 8**).

## DISCUSSION

Beyond confirming an inverse relationship between rare variants and the polygenic component of IPF risk, we evidenced in two IPF cohorts that models integrating IPF-PRS estimates improve the discriminatory power to stratify patients by the carrier status compared with models based on patient’s clinical history alone (22). The predictive ability of models integrating IPF-PRS estimates improved as more stringent criteria were applied for defining the carrier status and when patients were stratified by TL. These findings extend to IPF a principle previously described in other complex diseases (14) and suggest that IPF-PRS may serve as a complementary tool to prioritize rare variant discovery.

The genetic predisposition to IPF is governed by common variants of mild effect, by rare variants in telomere and non-telomere genes of large effect, and by short TL (3,23–25). The complex interplay between these factors remains incompletely understood, although the evidence suggests the existence of distinct genetic endotypes of IPF that should be considered in both research and clinical environments (11,26). In this context, in the patients showing low polygenic component, the relative contribution of the other genetic predictors is expected to be higher consistent with a liability threshold model of disease susceptibility (13,14). We previously explored this by assessing the correlation between IPF-PRS obtained from models based on sentinel variants, i.e., significant IPF risk variants in GWAS (11). Although results supported non-additive effects of rare and common variants, the association attenuated once the *MUC5B* variant was excluded from the IPF-PRS model. Here we demonstrate that IPF-PRS models incorporating additional SNPs beyond sentinel variants improves the robustness of this association. Importantly, excluding variants from the *MUC5B* locus did not alter the association. These results are consistent with previous studies highlighting the contribution of common variants other than the *MUC5B* region to disease risk (26,27).

Given this, we propose that integrating IPF-PRS along with clinical predictors has the potential to improve the identification of rare variant carriers. Family history of ILD and age of diagnosis strongly rely on patient recall, clinical interviews, and the physician expertise and are, therefore, prone to substantial bias. For example, relatives of patients with sporadic IPF have a high prevalence of undiagnosed ILD (28). This adds to a scenario where the disease often presents with non-specific symptoms, such as cough or dyspnoea, leading to diagnostic delays and uncertain assessment of the onset of disease (29). The limitation of these predictors is reflected in our results: in the PFF-PR the model based only on clinical history data had modest discrimination between carriers and non-carriers (AUC=0.62). This was more pronounced in the PROFILE cohort (AUC=0.58), where family history data was not available and age at diagnosis was not associated with carrier status. In contrast, the integration of IPF-PRS with the patient’s clinical history achieved a substantial improvement to predict the carriers. The improved performance was particularly notable for WG-PRS in the PFF-PR in comparison with Sentinel-PRS, which did not show significant differences with the model based only on clinical data. In this context, the Bayesian approach of IPF-PRS estimation demonstrated consistent results in an independent cohort, likely reflecting the improved performance by accurately weighting variants through the incorporation of functional annotations (20). One additional important feature to consider of this method is that it may allow the transferability of the model across populations of different ancestries (30).

The integration of WG-PRS or Sentinel-PRS with the clinical variables into a PRS model have shown the potential to identify additional carriers who would, otherwise, be missed following current standards (22). Early identification of carriers of monogenic variants, especially those affecting telomere genes, has important implications for clinical-decision making. First, these patients may require frequent monitoring of lung function, and early detection of extrapulmonary complications such as hematopoietic disorders or liver pathology (31,32). Second, this information could guide treatment decisions, especially in young patients, including adjusting of the immunosuppressive regime after lung transplantation to reduce haematological complications (33). Finally, identifying carriers may also justify screening of first-degree relatives for interstitial lung abnormalities or early ILDs, which may allow timely interventions and guidance to reduce environmental risk exposures (28,34). The improved detection of carriers, however, was accompanied by a higher rate of misclassification among non-carriers, which may raise concerns about the quality of the model (35). However, the clinical implications of such misclassification are limited since being classified as a carrier does not have the same clinical consequences as being a non-carrier. Therefore, in potential scenario of clinical integration of IPF-PRS estimates, a model that tends to overclassify potential carriers may be preferable. This is because gene sequencing itself is not associated with harm and the identification of carriers have implication for patient management and family screening, as previously described. Future studies should, however, assess the cost-effectiveness of extending gene sequencing to a larger number of IPF patients from the perspective of the healthcare system, as has been exposed in other genetic disorders (36,37).

Using alternative definitions of QVs confirmed a consistent direction in the performance of the IPF-PRS models across all definitions. Notably, the IPF-PRS models had better performance when more restrictive QVs criteria were applied. These results are consistent with our previous findings, where these other variant categories were associated with worse survival, further supporting their enrichment in deleteriousness potential (11). In addition, the models performed better when stratifying by TL indicating that they more accurately predict carriers of variants with functional consequences. These findings suggest that for a given carrier, the greater the penetrance of the rare variant, the smaller the relative contribution of PRS to disease risk, confirming the existence of distinct endotypes in IPF and the need for applying specific analytical approaches tailored to each of them. Taken together, IPF-PRS may be more useful for identifying individuals carrying highly penetrant deleterious variants, but would be less helpful for those carrying variants of incomplete penetrance or variable expressivity, where PRS may act as a genetic modifier, as suggested elsewhere (38).

This study has the following limitations. First, despite the growing interest in PRS and the potential clinical applications, they are not yet routinely implemented in clinical settings due to gaps in knowledge related to their use. Therefore genome-wide genotype data are not routinely available for IPF patients, whereas clinical variables are already embedded in established diagnostic and management frameworks for identifying rare variant carriers. While our findings contribute to a growing body of evidence on the interplay between common and rare genetic variants in IPF (26) the path from research utility to clinical implementation requires additional steps that go beyond the scope of this study. The scenario most immediately applicable for our findings is, therefore, the retrospective prioritization of sequencing in cohorts with existing SNP array data, rather than a replacement of established genetic testing workflows. Second, WG-PRS were estimated using WGS data. While WGS is increasingly adopted in IPF research cohorts, its use in the clinical practice is limited due to the high costs of the test. Low-coverage WGS may serve as a more cost-efficient alternative to provide PRS estimates (39,40), potentially enabling the sequential strategy proposed here. Future studies should therefore evaluate the concordance between WG-PRS and PRS derived from other more cost-effective approaches. Third, there is a lack of standards as to which is the best PRS method to use. Given their diversity, each with particular strengths and weaknesses,, the method of choice should provide a model that is transferable across genetic ancestries without compromising performance. To try to circumvent this limitation, here we have used three methods including two that allowed to extend the analyses beyond a model based just on the sentinel variants: PRsice-2 which uses the clumping and thresholding method and SBayesRC which uses a Bayesian approach incorporating with weights based on functional predictions. Future studies should evaluate the method which fits best in the genetic architecture of IPF (41). Fourth, the PRS performance is influenced by the availability of precise effect size estimates across different genetic ancestries. The GWAS used to derive SNP effect sizes had a limited sample size and was composed of patients of European genetic ancestry (4,125 IPF cases) (5). Future GWAS iterations involving larger and more diverse study sample may enhance the predictive power of IPF-PRS models. Fifth, QVs may not all be pathogenic since there is a lack of functional evidence or family segregation information in the study. Finally, TL estimates were derived from WGS from peripheral whole-blood DNA and were not age-adjusted. Although this is a promising approach, the method has not proved to outperform the gold standard (i.e., Flow-FISH) for clinical diagnosis (42).

## CONCLUSIONS

We confirm the non-additive effects between rare variants and common variants in IPF risk, supporting the existence of distinct endotypes. The integration of WG-PRS to clinical data models demonstrated improved discriminatory power for distinguishing carriers from non-carriers and enabled the identification of additional carriers beyond those detected using the patient’s clinical history alone. Further research is needed to evaluate the cost-effectiveness of incorporating PRS estimates into clinical workflows to identify patients that will likely benefit from gene sequencing.

## Data Availability

Data supporting the findings are available as part of the manuscript or from the supplementary files. Access to the raw whole-genome sequence dataset is restricted to qualified researchers under an agreement with the PFF-PR and PROFILE steering committees to protect the privacy of the participants. For further information and to apply for access to the data from the PFF-PR prior to public deposit in BioLINCC, please contact the chair of the ancillary committee (Dr. Noth) or any other member of the steering committee. For further information and to apply for access to the PROFILE data, please contact. Data access requests must be reviewed before release.

## LIST OF ABBREVIATIONS

IPF: idiopathic pulmonary fibrosis
GWAS: genome-wide association study
OR: odds ratio
NGS: next generation sequencing
PF: pulmonary fibrosis
RDVs: rare deleterious variants
PRS: polygenic risk score
WG: whole-genome
WGS: whole-genome sequencing
PFF-PR: Pulmonary Foundation Patients Registry
PROFILE: Prospective Observation of Fibrosis in the Lung Clinical Endpoints
QC: quality control
ILD: interstitial lung disease
TL: telomere length
QVs: qualifying variants
AF: allele frequency
SNPs: single nucleotide polymorphisms
PC: principal components
LD: linkage disequilibrium
AUC: area under the curve
ROC: receiver operating characteristic
NRI: net reclassification index
DCA: decision curve analysis
CI: confidence interval

## DECLARATIONS

### Ethics approval and consent to participate

Both studies (PFF-PR and PROFILE) were conducted according to The Declaration of Helsinki and written informed consent was obtained from all participants. Ethical approval for the PFF-PR was granted by the Institutional Review Board from the University of Michigan (MI, USA; ethics reference number HUM00111461). Ethical approval for the PROFILE study was granted by the Royal Free Hospital Research Ethics Committee (London, UK; ethics reference number 10/H0720/12) and University of Northampton Research Ethics Committee (Northampton, UK; ethics reference number 10/H0402/2).

### Consent for publication

Not applicable

### Author’s contributions

Conceptualization and supervision: A.A.G. and C.F. Patient recruitment, collection of biospecimen, and clinical data: J.M.O, A.A., F.J.M., T.M.M., R.G.J., I.N., S.F.M., E.S., J.M., J.S.K., Y.H., L.V.W., and R.J.A. Formal analysis: A.A.G., D.J., J.M.L.S., A.D., A.Q.B., B.G.G., D.Z., C.G., and D.Z. Supervision: I.N. and C.F. Writing-original draft: A.A.G. and C.F. Funding acquisition: C.F., L.V.W., A.A.G., M.M.M., and I.N. Visualization: A.A.G. Writing-review and editing: All the authors.

## Acknowledgements

We thank all patients who participated in the PFF Patient Registry. We also thank investigators and other staff at participating PFF Care Centers for providing clinical data and blood samples, the PFF which established and has maintained the Patient Registry since 2016, and lastly, the many generous donors.

---

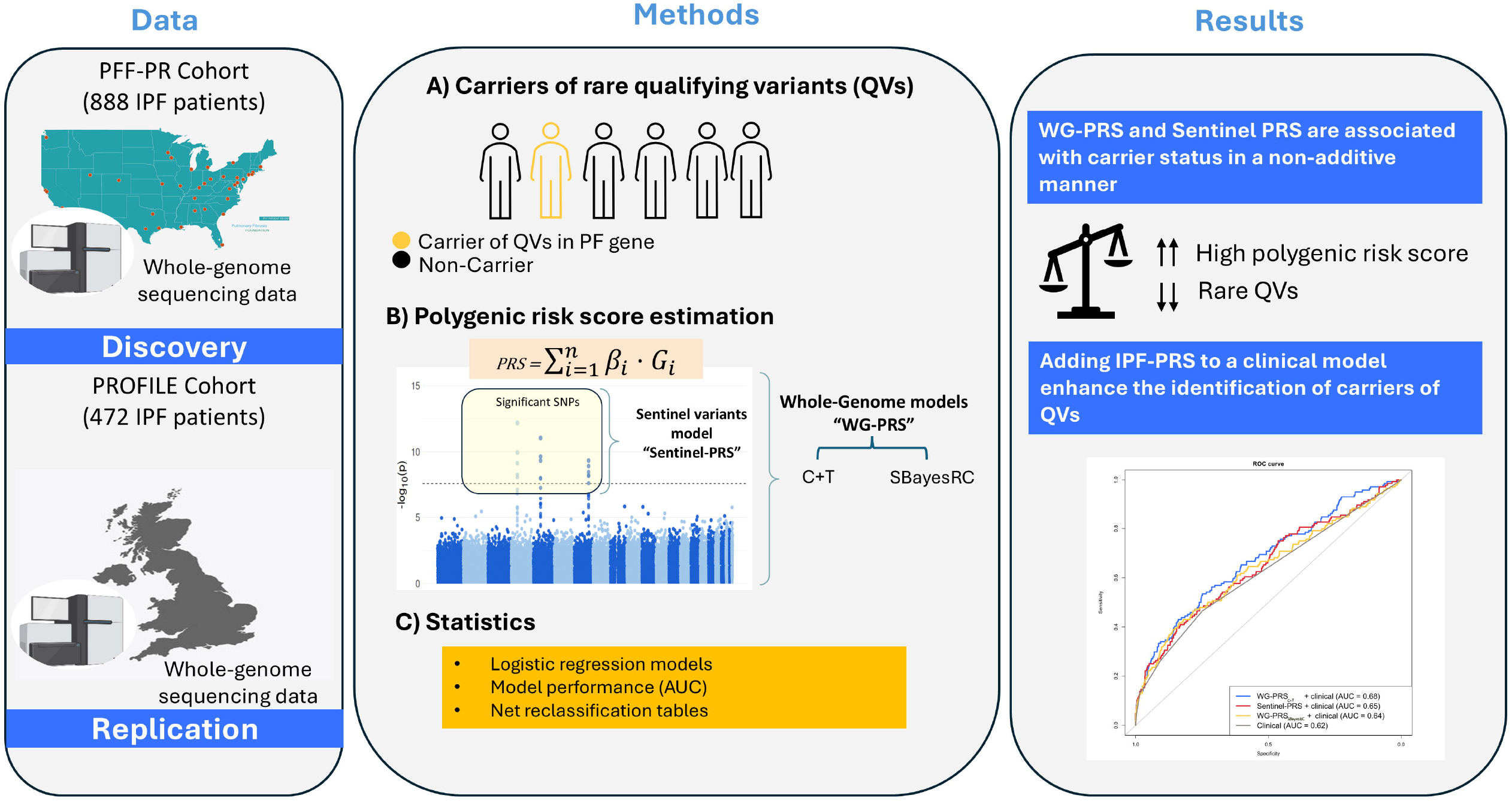

