## Additional file 1 for "Polygenic risk scores enhance the identification of carriers of monogenic forms of idiopathic pulmonary fibrosis"

**Supplementary Material**

| **Supplementary Table 1. Alternative definitions for qualifying variants and rare synonymous variants used for sensitivity analyses.** | | | | | |
| --- | --- | --- | --- | --- | --- |
|  | Telomere (QVs in telomere gene) | Ultra-rare Ensembl#  (PTV + Missense + Indel) | Rare  PTV only | Semi-rare Ensembl^#^  (PTV + Missense + Indel) | Rare synonymous^^^ |
| Missense AF* | 0.0005 | 0 | - | 0.01 | - |
| PTV AF* | 0.0005 | 0 | 0.001 | 0.01 | - |
| Consensus in silico prediction for missense:^&^ | | | | | |
| Polyphen2 Humdiv | - | Probably | - | Probably | - |
| REVEL | - | >0.5 | - | >0.5 | - |
| PrimateAI | - | >0.8 | - | >0.8 | - |
| CADD | >15 | - | - | - |  |
| Variants (n) | 105 | 30 | 28 | 78 | 38 |
| *Below threshold for any population in gnomAD v2.1 exomes (AFR, AMR, ASJ, EAS, FIN, NFE, OTH, SAS) or gnomAD v3.2 genomes (AFR, AMR, ASJ, EAS, FIN, MID, NFE, OTH, SAS) or in The 1000 Genomes Project Phase 3 genomes (AFR, AMR, EAS, EUR, SAS).  ^&^Consensus of three predictors (Polyphen2, REVEL, PrimateAI) for missense variants only if >2 out of 3, or 2 out of 2, or 1 out of 1 filters pass. Some predictors may have missing values.  ^^^Allele frequency cutoff of 0.0005 in any population in gnomAD v2.1 exomes (AFR, AMR, ASJ, EAS, FIN, NFE, OTH, SAS) or gnomAD v3.2 genomes (AFR, AMR, ASJ, EAS, FIN, MID, NFE, OTH, SAS) or in The 1000 Genomes Project Phase 3 genomes (AFR, AMR, EAS, EUR, SAS), only synonymous variants.  ^#^Ensembl models include non-coding *TERC* variants selected if passing missense AF level and involved in intramolecular base-pairing or previously described in pulmonary fibrosis or dyskeratosis congenita or Hoyeraal Hreidarsson syndrome.  PTV: Protein truncating variants. | | | | | |

| **Supplementary Table 2. Association of IPF-PRS with the carrier status in the PFF-PR idiopathic pulmonary fibrosis patients using alternative definitions of qualifying variants.** | | | | | |
| --- | --- | --- | --- | --- | --- |
|  | **OR (95% CI), p-value** | | | | |
| **Model** | **Telomere**  **(N=105)** | **Semi-rare Ensembl**  **(N=75)** | **Ultra-rare Ensembl**  **(N=30)** | **Rare PTV**  **(N=28)** | **Rare synonymous**  **(N=38)** |
| WG-PRS_C+T_ | 0.69 (0.55-0.85), p=8.41x10^-4^ | 0.59 (0.45-0.77), p=8.70x10^-5^ | 0.52 (0.34-0.77), p=1.43x10^-3^ | 0.57 (0.37-0.85), p=6.71x10^-3^ | 1.35 (0.97-1.88), p=0.072 |
| WG-PRS_SBayesRC_ | 0.68 (0.54-0.84),  p=4.82x10^-4^ | 0.57 (0.44-0.74), p=3.36x10^-5^ | 0.34 (0.21-0.53), p=3.04x10^-6^ | 0.47 (0.30-0.72), p=6.45x10^-4^ | 1.26 (0.91-1.75), p=0.161 |
| Sentinel-PRS | 0.75 (0.59-0.94), p=0.014 | 0.63 (0.47-0.83), p=1.35x10^-3^ | 0.39 (0.23-0.62), p=1.24x10^-4^ | 0.63 (0.39-0.97), p=0.041 | 1.18 (0.83-1.66), p=0.351 |
| IPF-PRS, polygenic risk score of idiopathic pulmonary fibrosis; N, number of carriers. OR, odds ratio; CI, confidence interval; PTV, protein truncating variants. | | | | | |

| Supplementary Table 3. Performance measures of models integrating IPF-PRS with patient’s clinical history in predicting the carrier status in the PFF-PR cohort of idiopathic pulmonary fibrosis patients. Models based only on patient’s clinical history are included for reference. | | | | | | | | | |
| --- | --- | --- | --- | --- | --- | --- | --- | --- | --- |
| Model | **Risk threshold**  **(%)** | **True positive** | **False negative** | **False positive** | **True negative** | **Sensitivity** | **Specificity** | **PPV** | **NPV** |
| WG-PRS_C+T_ + Clinical | 5 | 143 | 1 | 725 | 19 | 0.99 | 0.02 | 0.16 | 0.95 |
|  | 10 | 116 | 28 | 493 | 251 | 0.80 | 0.34 | 0.19 | 0.90 |
|  | 15 | 84 | 60 | 268 | 476 | 0.58 | 0.64 | 0.24 | 0.89 |
|  | 20 | 66 | 78 | 155 | 589 | 0.46 | 0.79 | 0.30 | 0.88 |
| WG-PRS_SBayesRC_ + Clinical | 5 | 144 | 0 | 738 | 6 | 1.00 | 0.008 | 0.16 | 1.00 |
|  | 10 | 121 | 23 | 519 | 225 | 0.84 | 0.30 | 0.19 | 0.91 |
|  | 15 | 87 | 57 | 256 | 488 | 0.60 | 0.65 | 0.25 | 0.89 |
|  | 20 | 62 | 82 | 147 | 597 | 0.43 | 0.80 | 0.30 | 0.88 |
| Sentinel-PRS + Clinical | 5 | 144 | 0 | 744 | 0 | 1.00 | 0 | 0.16 | NA |
|  | 10 | 122 | 22 | 559 | 185 | 0.85 | 0.25 | 0.18 | 0.89 |
|  | 15 | 76 | 68 | 239 | 505 | 0.53 | 0.68 | 0.24 | 0.88 |
|  | 20 | 63 | 81 | 157 | 587 | 0.44 | 0.79 | 0.29 | 0.88 |
| Clinical | 5 | 144 | 0 | 744 | 0 | 1.00 | 0 | 0.16 | NA |
|  | 10 | 144 | 0 | 744 | 0 | 1.00 | 0 | 0.16 | NA |
|  | 15 | 73 | 71 | 215 | 529 | 0.51 | 0.71 | 0.25 | 0.88 |
|  | 20 | 68 | 76 | 190 | 554 | 0.47 | 0.74 | 0.26 | 0.88 |
| IPF-PRS, polygenic risk score of idiopathic pulmonary fibrosis; PPV, positive predictive value; NPV, negative predictive value; NA, not available. | | | | | | | | | |

| Supplementary Table 4. Performance measures of models integrating IPF-PRS with patient’s clinical history in predicting the carrier status in the PROFILE cohort of idiopathic pulmonary fibrosis patients. Models based only on patient’s clinical history are included for reference. | | | | | | | | | |
| --- | --- | --- | --- | --- | --- | --- | --- | --- | --- |
| Model | **Risk threshold**  **(%)** | **True positive** | **False negative** | **False positive** | **True negative** | **Sensitivity** | **Specificity** | **PPV** | **NPV** |
| Sentinel-PRS + Clinical | 5 | 51 | 1 | 392 | 28 | 0.98 | 0.06 | 0.11 | 0.96 |
|  | 10 | 28 | 24 | 185 | 235 | 0.53 | 0.55 | 0.13 | 0.90 |
|  | 15 | 17 | 62 | 35 | 358 | 0.33 | 0.85 | 0.21 | 0.91 |
|  | 20 | 10 | 42 | 20 | 400 | 0.20 | 0.95 | 0.33 | 0.90 |
| WG-PRS_SBayesRC_ + Clinical | 5 | 74 | 0 | 396 | 2 | 0.94 | 0.05 | 0.11 | 0.88 |
|  | 10 | 41 | 33 | 173 | 225 | 0.60 | 0.55 | 0.14 | 0.92 |
|  | 15 | 14 | 60 | 47 | 351 | 0.30 | 0.85 | 0.20 | 0.91 |
|  | 20 | 14 | 60 | 47 | 351 | 0.23 | 0.95 | 0.36 | 0.91 |
| Clinical | 5 | 52 | 0 | 418 | 2 | 1 | 0.004 | 0.11 | 1 |
|  | 10 | 32 | 20 | 182 | 238 | 0.61 | 0.56 | 0.14 | 0.92 |
|  | 15 | 12 | 40 | 49 | 371 | 0.23 | 0.88 | 0.20 | 0.90 |
|  | 20 | 7 | 45 | 20 | 400 | 0.13 | 0.95 | 0.25 | 0.89 |
| IPF-PRS, polygenic risk score of idiopathic pulmonary fibrosis; PPV, positive predictive value; NPV, negative predictive value; NA, not available. | | | | | | | | | |

| **Supplementary Table 5. Association of IPF-PRS with the qualifying variant carrier status and the area under the curve (AUC) for the carrier prediction with different models and stratifying by clinical variables in the PFF-PR cohort of patients with idiopathic pulmonary fibrosis.** DeLong’s test p-values are indicated reflecting the performance comparison of models integrating the IPF-PRS with the patient’s clinical history (integrative) vs. models based only on the patient’s clinical history (clinical). | | | | | | | |
| --- | --- | --- | --- | --- | --- | --- | --- |
| **Model** | **Variable** | **N** | **Covariates** | **OR**  **(95% CI), p** | **AUC**  **(Integrative)** | **AUC**  **(Clinical)** | **DeLong’s Test p-value** |
| **WG-PRS_C+T_** | AaD <50 y.o. | 22 | FPF, TL≤10th, PC1-2 | 0.25 (0.02-1.01), 0.14 | 0.90 | 0.79 | 0.13 |
|  | AaD $\geq$50 y.o. | 866 | FPF, TL≤10th, PC1-2 | 0.66 (0.54-0.81), 4.80x10^-5^ | 0.66 | 0.61 | 0.02 |
|  | TL ≤10^th^ | 89 | Age<50, FPF, PC1-2 | 0.45 (0.24-0.76), 6.0x10^-3^ | 0.75 | 0.62 | 0.04 |
|  | TL >10^th^ | 799 | Age<50, FPF, PC1-2 | 0.69 (0.56-0.85), 5.46x10^-3^ | 0.63 | 0.58 | 0.02 |
|  | Family history | 176 | Age<50, TL≤10th, PC1-2 | 0.55 (0.35-0.81), 3.81x10^-3^ | 0.75 | 0.65 | 0.02 |
|  | Sporadic | 712 | Age<50, TL≤10th, PC1-2 | 0.69 (0.55-0.86), 1.23x10^-3^ | 0.62 | 0.57 | 0.11 |
| **WG-PRS_SBayesRC_** | AaD <50 y.o. | 22 | FPF, TL≤10th, PC1-2 | 0.49 (0.13-1.31), 0.20 | 0.88 | 0.79 | 0.30 |
|  | AaD $\geq$50 y.o. | 866 | FPF, TL≤10th, PC1-2 | 0.71 (0.59-0.87), 7.22x10^-4^ | 0.65 | 0.61 | 0.14 |
|  | TL≤10^th^ | 89 | Age<50, FPF, PC1-2 | 0.43 (0.23-0.75), 5.15x10^-3^ | 0.74 | 0.63 | 0.08 |
|  | TL >10^th^ | 799 | Age<50, FPF, PC1-2 | 0.76 (0.62-0.94), 9.34x10^-3^ | 0.61 | 0.57 | 0.11 |
|  | Family history | 176 | Age<50, TL≤10th, PC1-2 | 0.65 (0.44-0.92), 0.02 | 0.73 | 0.66 | 0.05 |
|  | Sporadic | 712 | Age<50, TL≤10th, PC1-2 | 0.73 (0.58-0.92), 7.86x10^-3^ | 0.60 | 0.57 | 0.28 |
| **Sentinel-PRS** | AaD <50 y.o. | 22 | FPF, TL≤10th, PC1-2 | 0.42 (0.07-1.51), 0.23 | 0.88 | 0.79 | 0.30 |
|  | AaD $\geq$50 y.o. | 866 | FPF, TL≤10th, PC1-2 | 0.78 (0.63-0.96), 0.02 | 0.63 | 0.61 | 0.31 |
|  | TL≤10^th^ | 89 | Age<50, FPF, PC1-2 | 0.42 (0.21-0.78), 9.39x10^-3^ | 0.74 | 0.62 | 0.06 |
|  | TL >10^th^ | 799 | Age<50, FPF, PC1-2 | 0.84 (0.67-1.04), 0.11 | 0.60 | 0.57 | 0.25 |
|  | Family history | 176 | Age<50, TL≤10th, PC1-2 | 0.73 (0.49-1.07), 0.11 | 0.71 | 0.66 | 0.12 |
|  | Sporadic | 712 | Age<50, TL≤10th, PC1-2 | 0.79 (0.62-1.00), 0.05 | 0.58 | 0.56 | 0.57 |

IPF-PRS, polygenic risk score of idiopathic pulmonary fibrosis; AaD, age at diagnosis; OR, odds ratio; CI, confidence interval; TL, telomere length; PC1-2, principal components 1 and 2.

| **Supplementary Table 6. Association of IPF-PRS with the qualifying variant carrier status and the area under the curve (AUC) for the carrier prediction with different models and stratifying by clinical variables in the PROFILE cohort of patients with idiopathic pulmonary fibrosis.** DeLong’s test p-values are indicated reflecting the performance comparison of models integrating the IPF-PRS with the patient’s clinical history (integrative) vs. models based only on the patient’s clinical history (clinical). | | | | | | | |
| --- | --- | --- | --- | --- | --- | --- | --- |
| **Model** | **Variable** | **N** | **Covariates** | **OR (95% CI), p** | **Integrative** | **Clinical** | **DeLong’s Test p-value** |
| **Sentinel-PRS** | AaD <50 y.o. | 4 | FPF, TL<10th, PC1-2 | 1.00  (0.00-inf), >0.9 | - | - | - |
|  | AaD $\geq$50 y.o. | 468 | FPF, TL<10th, PC1-2 | 0.68  (0.50-0.93), 0.02 | 0.62 | 0.58 | 0.31 |
|  | TL≤10^th^ | 48 | Age<50, FPF, PC1-2 | 0.20  (0.05-0.55), 6.27x10^-3^ | 0.93 | 0.66 | 0.03 |
|  | TL>10^th^ | 424 | Age<50, FPF, PC1-2 | 0.80  (0.57-1.11), 0.20 | 0.57 | 0.55 | 0.68 |
| **WG-PRS_SBayesRC_** | AaD <50 y.o. | 4 | FPF, TL<10th, PC1-2 | 1.00  (0.00-inf), >0.9 | - | - | - |
|  | AaD $\geq$50 y.o. | 468 | FPF, TL<10th, PC1-2 | 0.69  (0.50-0.93), 0.02 | 0.62 | 0.58 | 0.30 |
|  | TL≤10^th^ | 48 | Age<50, FPF, PC1-2 | 0.30  (0.09-0.74), 0.02 | 0.91 | 0.66 | 0.04 |
|  | TL >10^th^ | 424 | Age<50, FPF, PC1-2 | 0.78  (0.55-1.08), 0.13 | 0.58 | 0.55 | 0.64 |
| IPF-PRS, polygenic risk score of idiopathic pulmonary fibrosis; AaD, age at diagnosis; OR, odds ratio; CI, confidence interval; TL, telomere length; FPF, familial history of interstitial lung disease; PC1-2, principal components 1 and 2. | | | | | | | |

| **Supplementary Table 7. Area under the curve (AUC) for models predicting the carrier status in idiopathic pulmonary fibrosis patients from the PFF-PR using alternative definitions of qualifying variants.** DeLong’s test p-values are indicated reflecting the performance comparison of models integrating the IPF-PRS with the patient’s clinical history (integrative) *vs.* models based only on the patient’s clinical history (clinical). | | | | | | | | | | | | | | | |
| --- | --- | --- | --- | --- | --- | --- | --- | --- | --- | --- | --- | --- | --- | --- | --- |
| **Model** | **Telomere**  **(N=105)** | | | **Semi-rare Ensemble**  **(N=75)** | | | **Ultra-rare Ensemble**  **(N=30)** | | | **Rare PTV**  **(N=28)** | | | **Rare synonymous**  **(N=38)** | | |
|  | Integrative | Clinical | p-value | Integrative | Clinical | p-value | Integrative | Clinical | p-value | Integrative | Clinical | p-value | Integrative | Clinical | p-value |
| WG-PRS_C+T_ | 0.69 | 0.64 | 0.01 | 0.71 | 0.63 | 2.12x10^-3^ | 0.73 | 0.64 | 5.98x10^-3^ | 0.72 | 0.62 | 8.10x10^-3^ | 0.62 | 0.52 | 0.02 |
| WG-PRS_SBayesRC_ | 0.68 | 0.64 | 0.03 | 0.71 | 0.63 | 3.57x10^-3^ | 0.80 | 0.64 | 2.75x10^-3^ | 0.72 | 0.62 | 0.05 | 0.61 | 0.52 | 0.05 |
| Sentinel-PRS | 0.68 | 0.64 | 0.06 | 0.69 | 0.63 | 0.01 | 0.77 | 0.64 | 0.01 | 0.68 | 0.62 | 0.10 | 0.59 | 0.52 | 0.07 |

IPF-PRS, polygenic risk score of idiopathic pulmonary fibrosis; N, number of carriers; PTV, protein truncating variants.

| **Supplementary Table 8. Decision curve analysis.** Net benefit for each model at thresholds 5%, 10%, 15% and 20%. | | | | |
| --- | --- | --- | --- | --- |
| **Risk threshold (%)** | **Clinical** | **WG-PRS_C+T_**  **+**  **clinical** | **WG-PRS_SBayesRC_**  **+**  **clinical** | **Sentinel-PRS**  **+**  **clinical** |
| 0 | 0.16 | 0.16 | 0.16 | 0.16 |
| 5 | 0.12 | 0.12 | 0.12 | 0.12 |
| 10 | 0.07 | 0.07 | 0.07 | 0.07 |
| 15 | 0.04 | 0.04 | 0.05 | 0.04 |
| 20 | 0.02 | 0.03 | 0.03 | 0.03 |

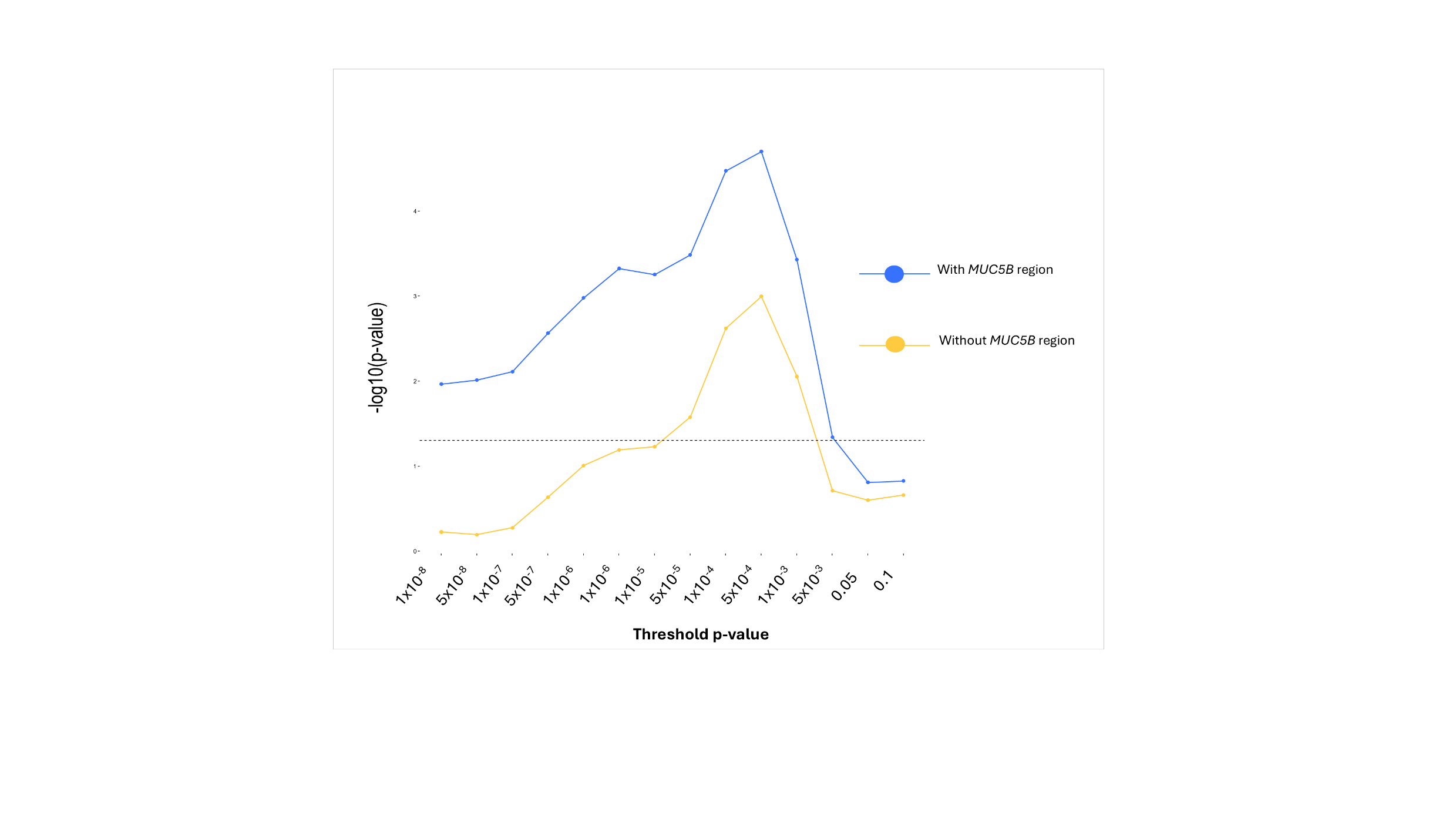

**Supplementary Figure 1. WG-PRS_C+T_ model-fitting and strength of association with the carrier status using logistic regression models.** The y-axis shows the negative log_10_ of the p-values estimated using logistic regression models. The x-axis indicates the p-value thresholds used to derive each WG-PRS. The dashed line represents the nominal significance threshold. The blue line shows the results from models including the MUC5B region, while the yellow line represents the results from models excluding the MUC5B region.

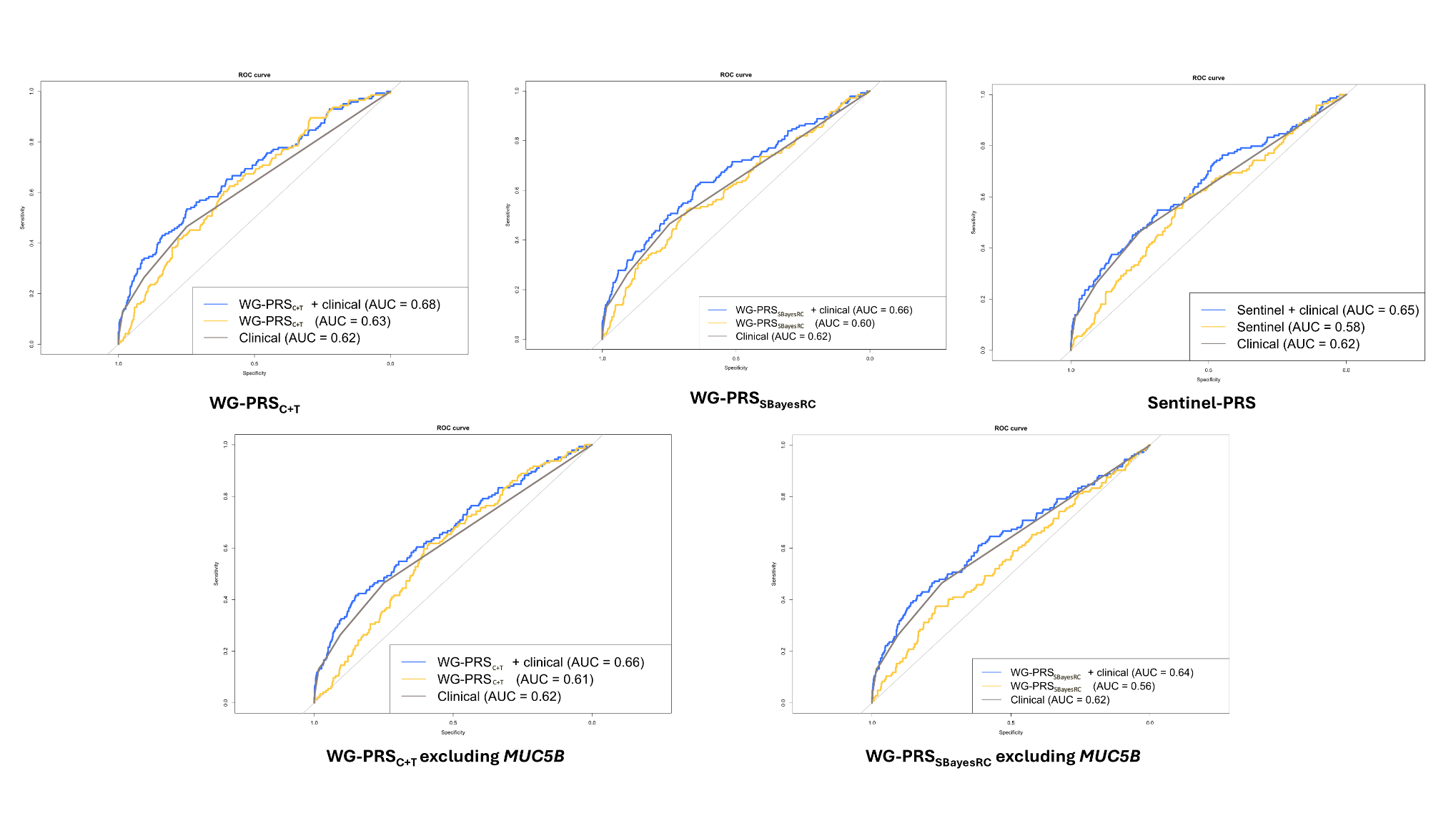
**Supplementary Figure 2.** Receiver operating characteristic (ROC) curves for carrier prediction in IPF patients from the PFF-PR based on a model with patient’s clinical history alone (age of diagnosis, family history, and telomere length), IPF-PRS estimates only, and a model integrating IPF-PRS and the patient’s clinical history.

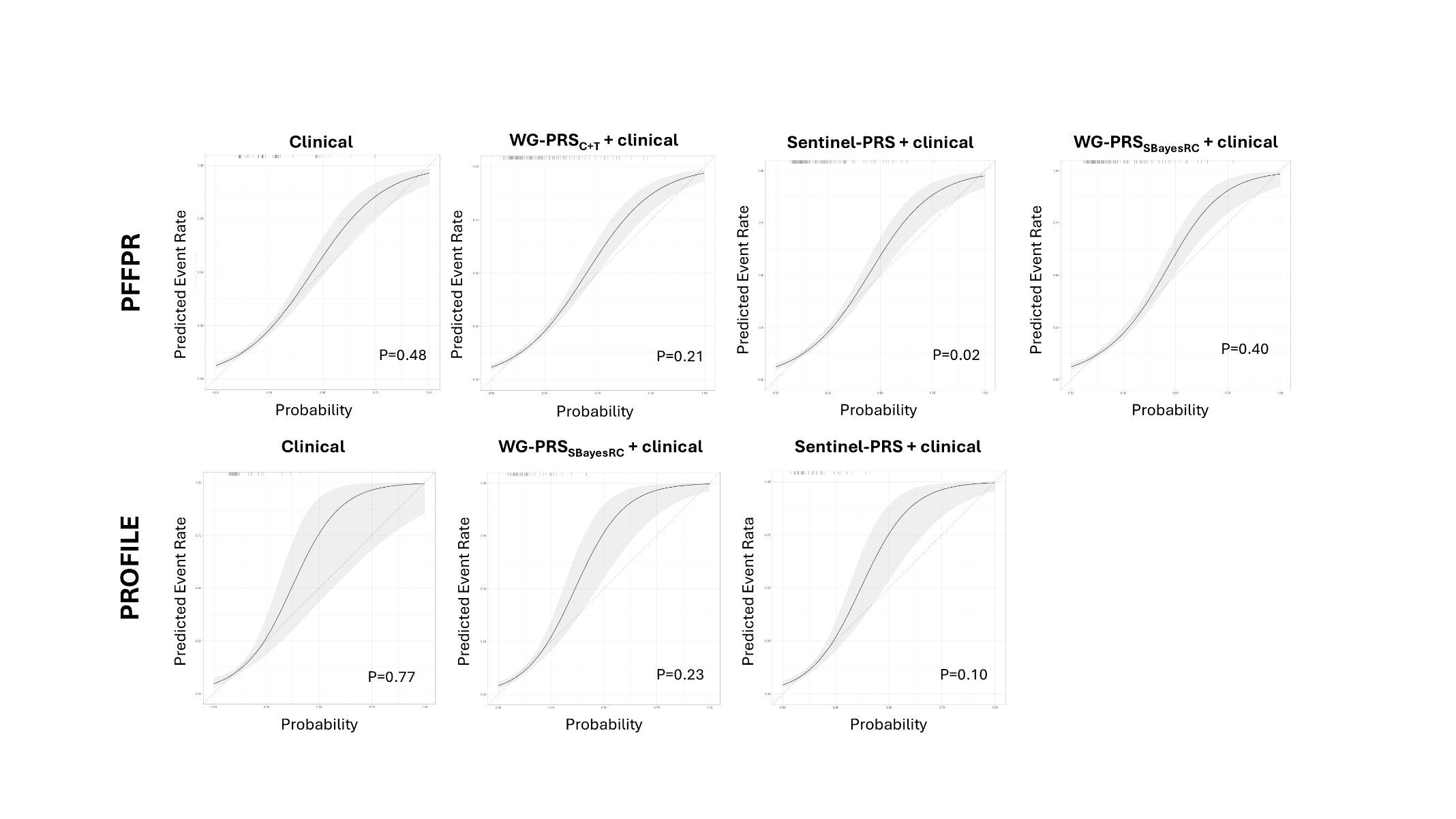

**Supplementary Figure 3.** Observed and predicted values of a model integrating the IPF-PRS with the patient’s clinical history and a model with the patient’s clinical history alone. The grey shading represents the confidence interval. Hosmer-Lemeshow p-values are shown.

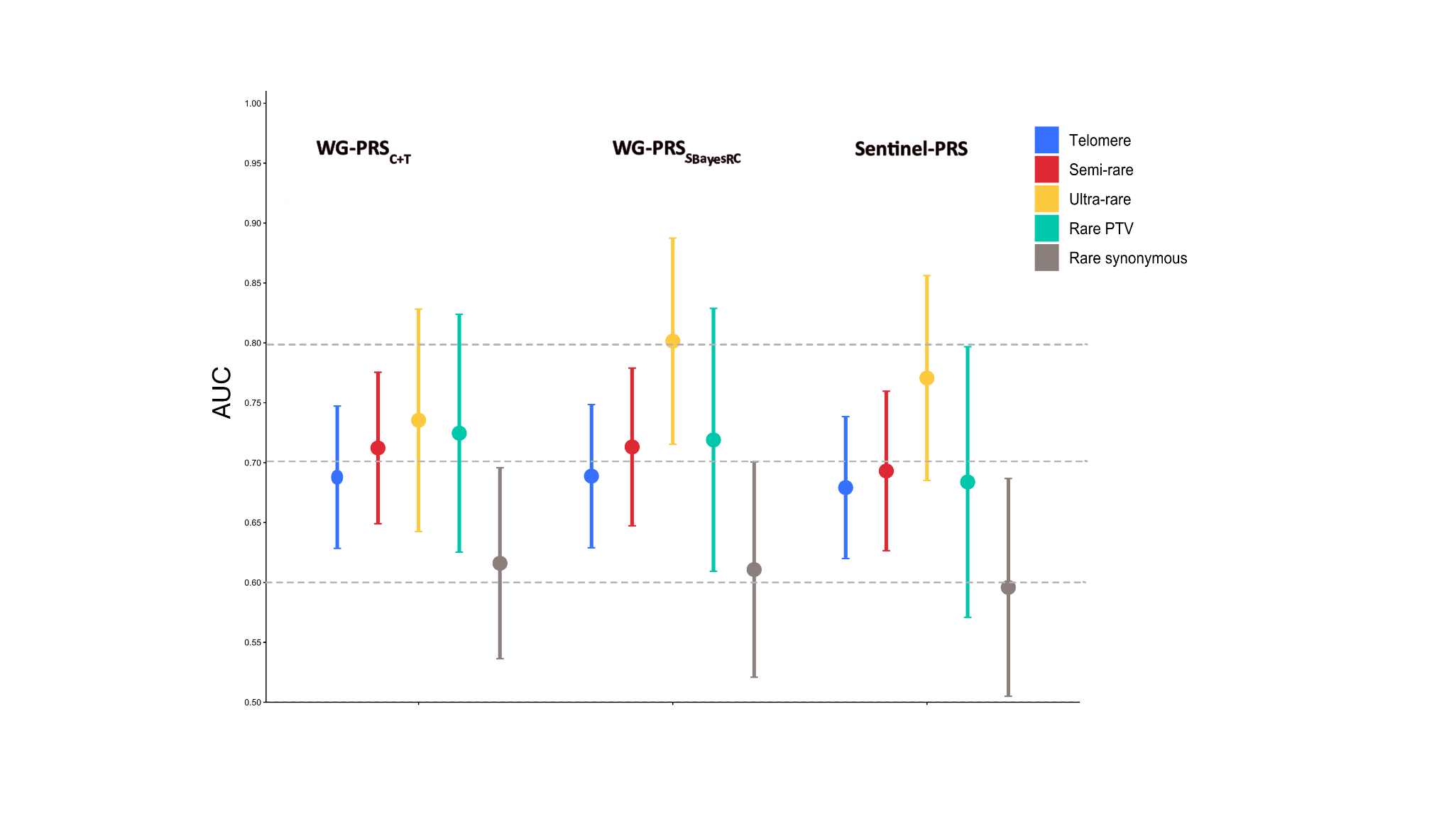

**Supplementary Figure 4.** Area under the curve (AUC) for predicting carrier status with models integrating the IPF-PRS plus the patient’s clinical history using alternative definitions of qualifying variants.

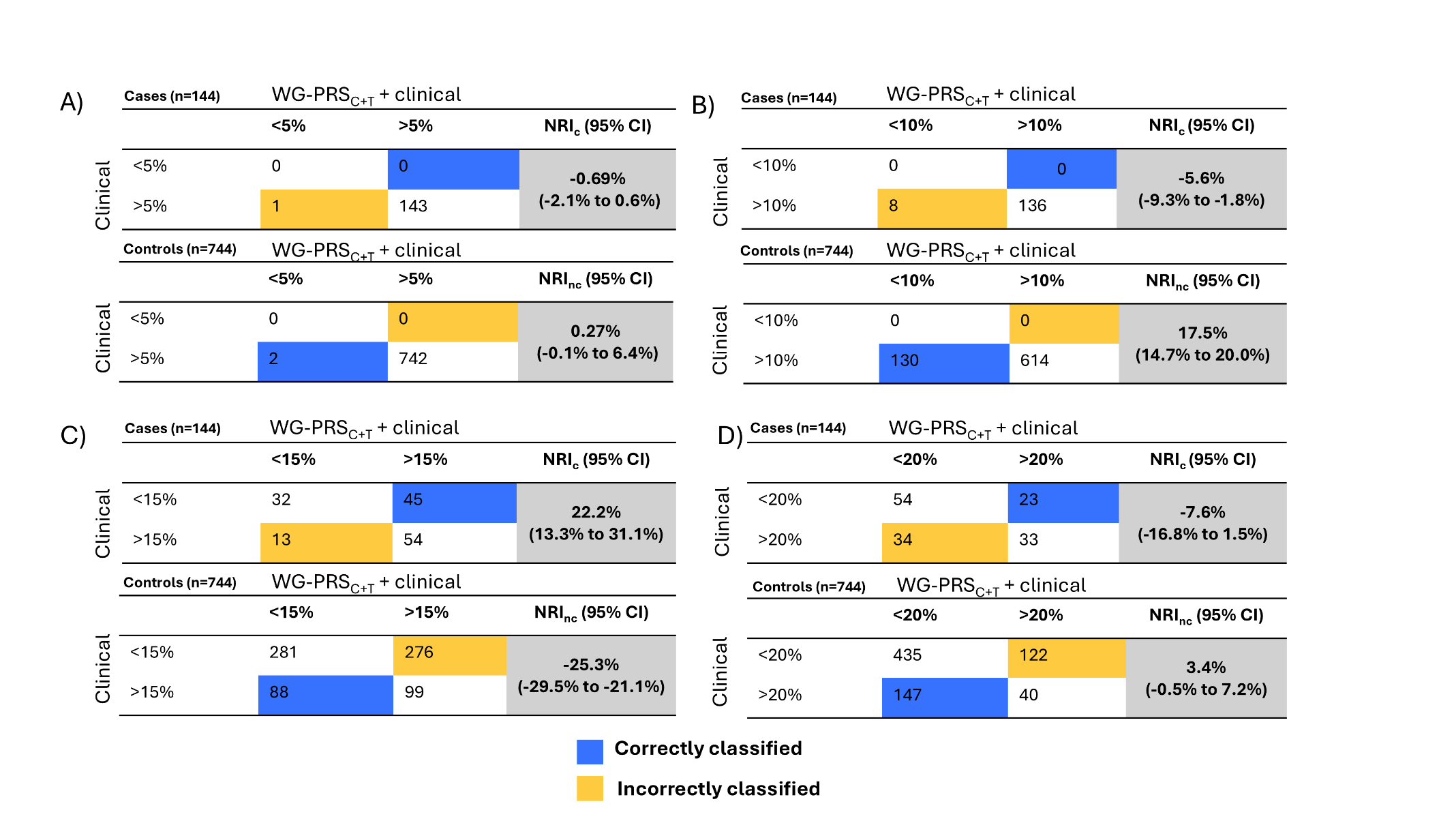

**Supplementary Figure 5.** Net Reclassification Tables comparing models integrating WG-PRS_C+T_ with the patient’s clinical data vs. the clinical model alone in the PFF-PR. The net reclassification index (NRI) quantifies how well the model (WG-PRS_C+T_ + clinical) correctly reclassifies individuals compared to an older model (clinical). CI=confidence interval; NRI_c_ for cases and NRI_nc_ for non-cases.

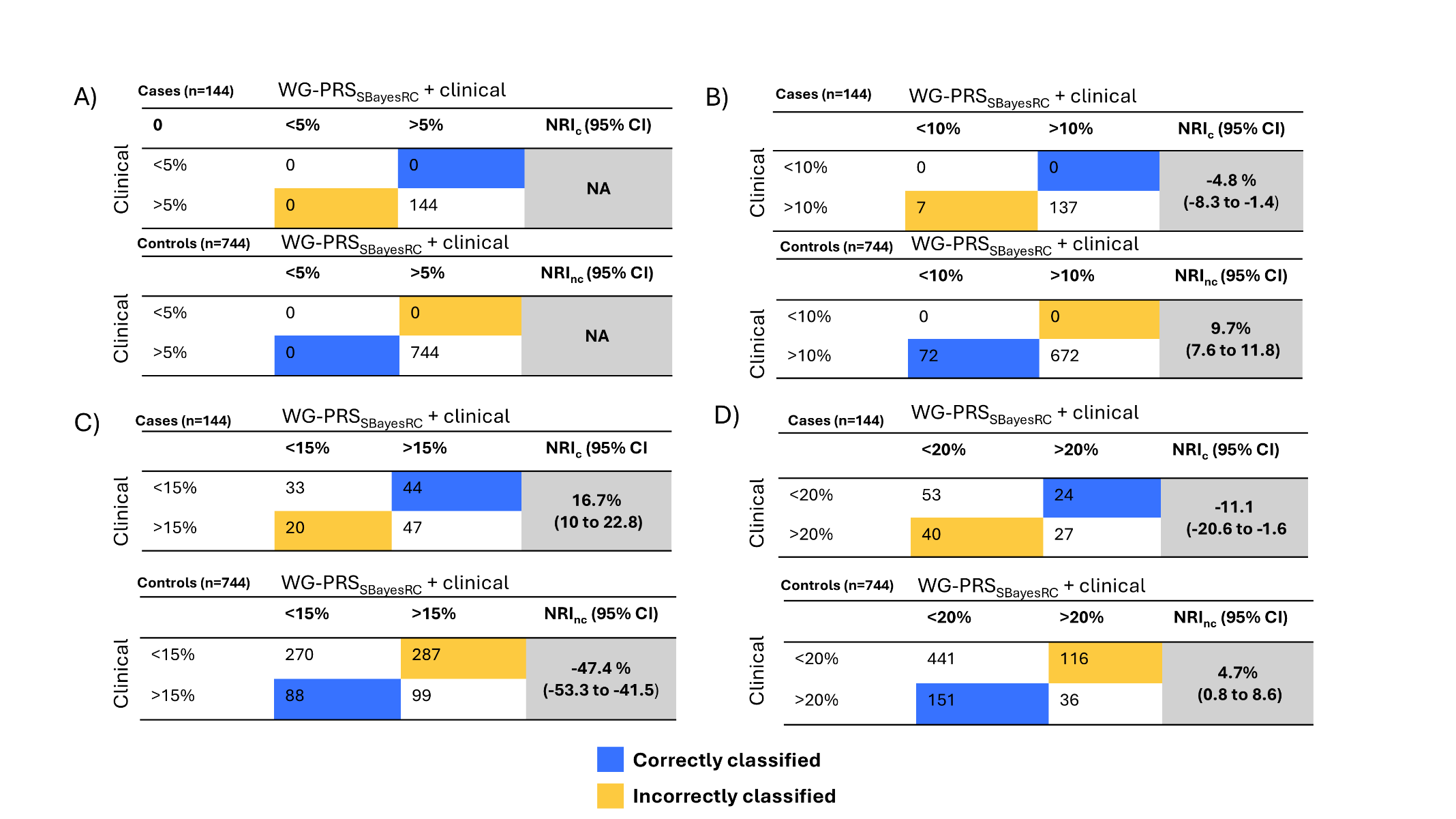

**Supplementary Figure 6.** Net Reclassification Tables comparing models integrating WG-PRS_SBayesRC_ with the patient’s clinical data vs. the clinical model alone in the PFF-PR. The net reclassification index (NRI) quantifies how well the model (WG-PRS_SBayesRC_+ clinical) correctly reclassifies individuals compared to an older model (clinical). CI=confidence interval; NRIc for cases and NRInc for non-cases.

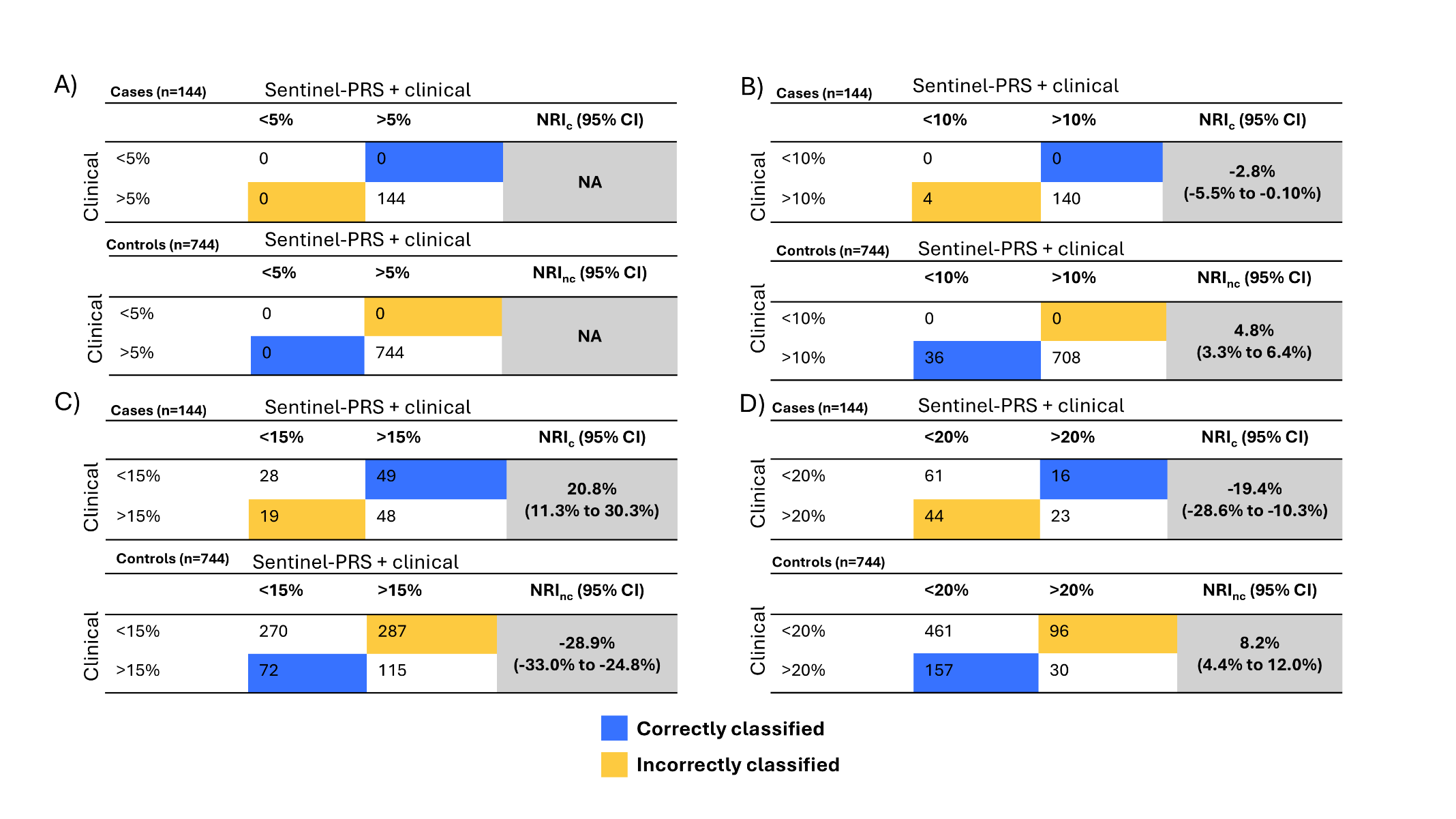

**Supplementary Figure 7.** Net Reclassification Tables comparing models integrating Sentinel-PRS with the patient’s clinical data vs. the clinical model alone in the PFF-PR. The net reclassification index (NRI) quantifies how well the model (Sentinel-PRS + clinical) correctly reclassifies individuals compared to an older model (clinical). CI=confidence interval; NRI_c_ for cases and NRI_nc_ for non-cases.

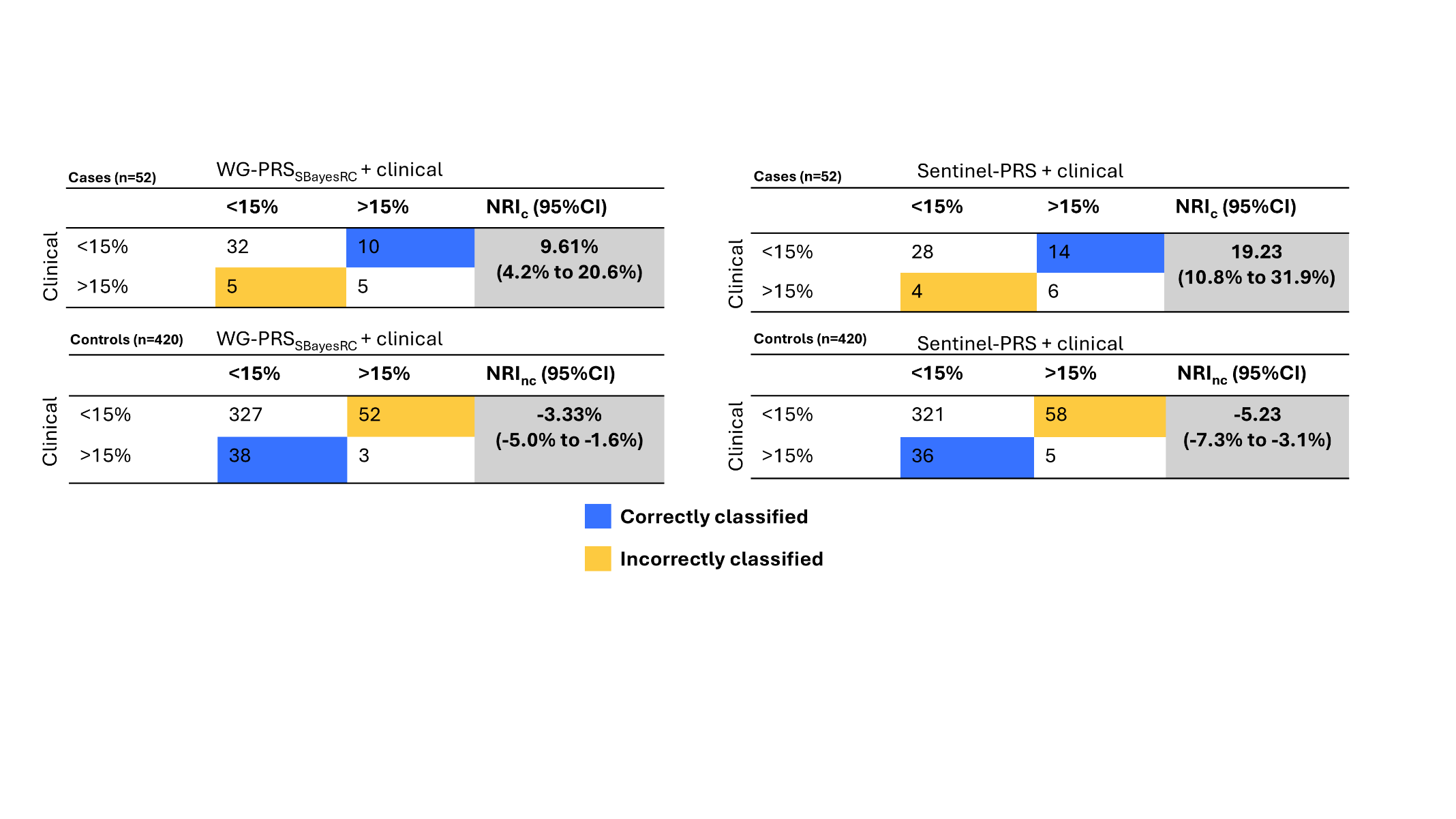

**Supplementary Figure 8.** Net Reclassification Tables comparing models integrating Sentinel-PRS or WG-PRSS_BayesRC_ with the patient’s clinical data vs. the clinical model alone in PROFILE. The net reclassification index (NRI) quantifies how well the model (Sentinel-PRS + clinical or WG-PRSS_BayesRC_ + clinical ) correctly reclassifies individuals compared to an older model (clinical). CI=confidence Interval; NRI_c_ for cases and NRI_nc_ for non-cases.

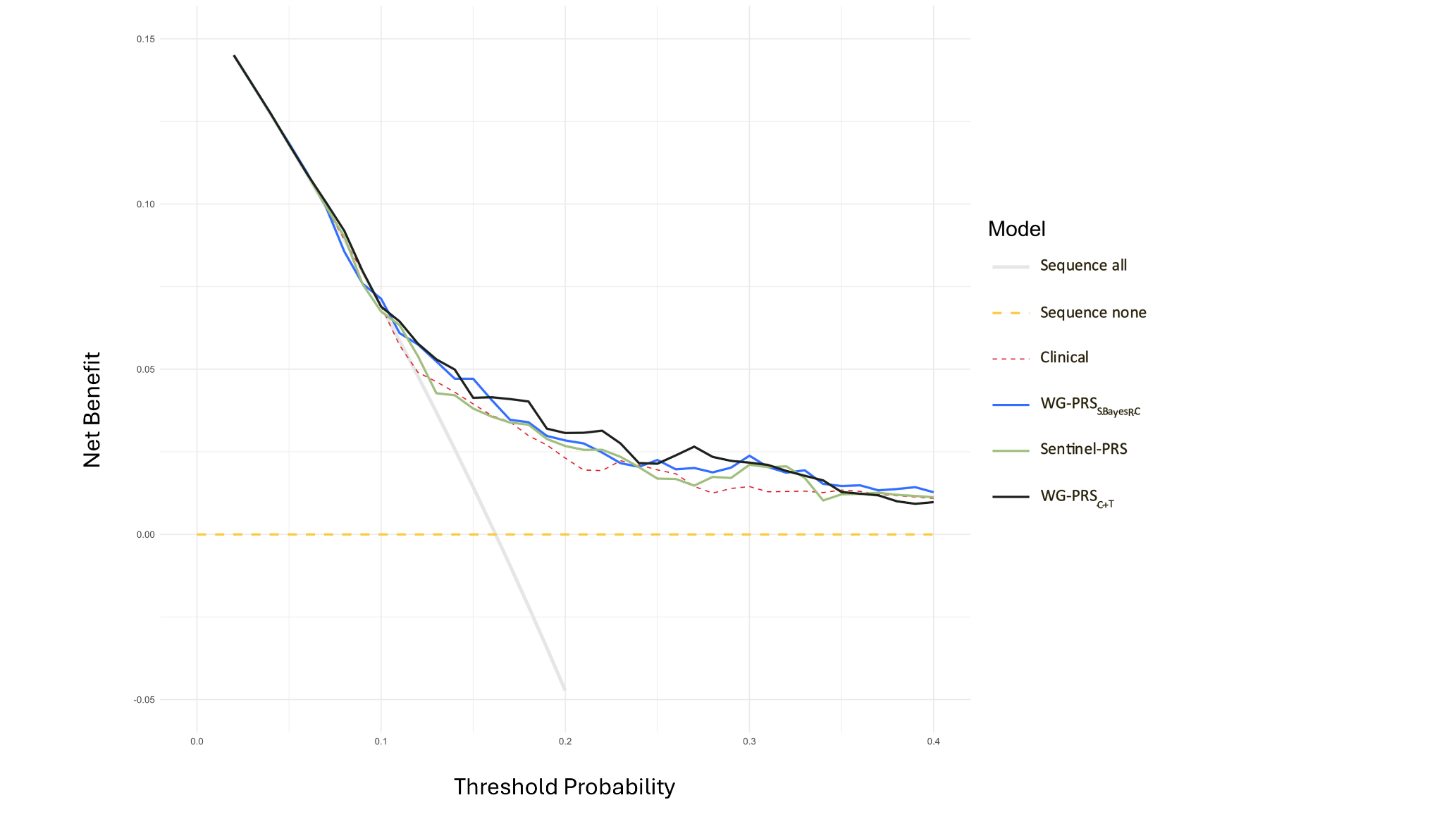
**Supplementary Figure 9**. Decision curve analysis plot. The clinical and IPF-PRS + clinical models are compared to the two default strategies of “Sequence All” and “Sequence none”.
